# HEALTH-SEEKING BEHAVIOR AND INTERPERSONAL INFLUENCES AMONG HYPERTENSIVE PATIENTS IN AN URBAN COMMUNITY

**DOI:** 10.1101/2025.06.12.25329475

**Authors:** Arbeen A. Laurito, Maria Graziela Louise M. Tomboc, Thessa Riz J. Paulo, Kent Lawrence K. Quitoriano, Martin Wyne P. Tion, Zamantha Alexzandra Marie E. Torregosa, Krisha Jane A. Villareal, Sofia G. Waminal

## Abstract

Hypertension is a prevalent chronic condition among Filipinos and is one of the leading causes of mortality, as it does not commonly manifest itself. The cornerstone of recommended care for hypertension is early detection, lifestyle changes, and medication adherence. However, adopting health-promoting behaviors is not as easy as it seems, for it is affected by one’s social context and health beliefs. This paper aimed to determine health-seeking Behavior and interpersonal influences, those resulting from one’s norms, social support, and modeling, as theorized by Pender. This study utilized the descriptive research design, participated by 60 hypertensive residents from an urban barangay in Cagayan de Oro City, Philippines using a modified survey questionnaire from The Interpersonal Support Evaluation List (ISEL) for gathering data and descriptive statistics as statistical techniques. Results yielded that the majority of the participants belonged to the age group of 65 and above, were female, and married. Seeking consultation is the most practiced health-seeking Behavior, followed by self-medication, lifestyle modification, and the least was alternative management. Norms and social support were very influential in managing hypertension, while modeling was only somewhat influential. Enforcing health-seeking Behavior through setting expectations and providing support can be effective in managing the disease. This study promotes awareness that the direct involvement of significant others in caring for hypertension may efficiently produce positive outcomes and significantly reduce mortality.

## Chapter 1 THE PROBLEM AND ITS SCOPE

### Introduction

Hypertension, or high blood pressure, is one of the leading causes of mortality and is also considered the leading preventable risk factor for cardiovascular disease. It continues to be a major public health problem globally, being called the “Silent killer,” due to its absence of obvious symptoms. It is estimated that 1.3 billion people are affected worldwide (World Heart Federation, n.d.). In the Philippine setting, hypertension affects 466,383 people nationally, and 45,425 locally in Cagayan de Oro City (Department of Health [DOH], 2019). According to Sakboonyarat et al. (2019), timely diagnosis and appropriate treatment are essential in controlling hypertension and preventing its severe complications such as cardiovascular diseases, stroke, or kidney failure.

Irwan et al. (2022) indicated that lifestyle modifications in diet, exercise, and stress management are effective to reduce the incidence of and improve management of hypertension. When combined with improved adherence to medications, these behaviors represent a cornerstone of recommended care for hypertension care (Carey et al., 2019). However, it should be noted that changing health behaviors is not easy as it requires extensive efforts and motivations from each individual, as influenced by one’s social context, which poses critical importance for their health enhancement (Giena et al., 2018). Carey et al. (2019) explained that interventions may fail ultimately due to the following reasons: (1) Patients are not well-adapted to the organization where implementation occurs; (2) Clinicians do not endorse the intervention; (3) Patients and their families are not actively engaged; and, (4) The community that patients live faces many challenges in implementing interventions. Heinert (2019) stated that the most common facilitators for hypertension control were social support, knowing how to control hypertension, and community resources. Any shortcomings in these aspects pose a threat to a patient’s management of hypertension.

Knowing the concept of health-seeking behavior is pertinent in understanding the factors that influence a certain patient’s disease management. By definition, it refers to any activity undertaken by individuals who perceive themselves to have a health problem or are in a state of illness to find an appropriate remedy (Gupta et al., 2020). Meanwhile, the factors that can influence hypertension awareness and control are interpersonal relationships and community resources (Pirkle et al., 2018). Smooth interpersonal relationships, as stated by Stanford Medicine (n.d.), are a core value for every community.

Despite the investigation of different studies on the predictors of health behaviors among individuals with hypertension, health-seeking behaviors and interpersonal influences among Filipinos with the said disease have yet to be studied further. Investigating an individual’s interpersonal relationships as the root for adopting health-seeking behaviors will also provide significant findings as to how respective Filipino households and communities play a role in hypertension management. This paper explored the health-seeking behaviors adopted by hypertensive patients in terms of the three interpersonal influences; Norms (family), peers (support), and models (providers). The study has investigated the participant’s demographic profile, health-seeking behavior, and interpersonal influences.

#### Theoretical and Conceptual Framework

The study is anchored on the health promotion model (HPM), proposed by Nola J. Pender in 1982, and revised in 1996. Health-promoting behavior was defined by Pender as an endpoint or action outcome that is directed toward attaining positive health outcomes such as optimal well-being, personal fulfillment, and productive living.

Moreover, it is asserted in the first book, Health Promotion in Nursing Practice, that individuals are motivated by complex biopsychosocial processes to engage in behaviors directed toward the enhancement of health (Alligood, 2017). Pender’s Health Promotion Model is one of the most widely used models to identify and change unhealthy behaviors, in turn, promoting health (Chen & Hsieh, 2021).

As stated by LaMorte (2022), the two components of health-related behavior are: (1) The desire to avoid illness, or conversely achieve wellness if already ill; and, (2) The belief that a specific health action will prevent, or cure, illness. To discuss the foundations of the Health Promotion Model, Khodaveisi et al., (2017) explained that it is based on social cognitive theory where cognitive-perceptual factors (perceived benefits, barriers, and self-efficacy) influence an individual’s engagement in health-promoting behaviors. Furthermore, modifying factors (Demographic characteristics, interpersonal influences, and behavioral factors) are known to interact with each other which in turn influences cognitive perceptual processes. This study mainly focuses on the variables mentioned in the latter statement. The investigator aims to determine the health-promoting behavior and interpersonal influences among hypertensive patients.

The theory supports that personal factors and prior related behavior contribute to the interpersonal influencers on an individual’s health-promoting behavior. Furthermore, three of the fourteen theoretical assertions derived from the model in the fourth edition of the book have shown specific relevance to this study. These assertions indicate that when significant others model the behavior, expect the behavior to occur, and provide assistance and support to enable the behavior, persons are more likely to commit to and engage in health-promoting behaviors. Next, important sources of interpersonal influences include families, peers, and healthcare providers. They can increase or decrease commitment to and engagement in health-promoting behavior. Lastly, to create incentives for health actions, people can modify cognitions, affect, and interpersonal and physical environments (Murdaugh et al., 2018).

This model is relevant to the study as it shows the interplay of how individual characteristics and experiences contribute to behavior-specific cognitions and affect, which then influences behavioral outcomes. To further support this, Gorbani et al. (2022) revealed that the Health Promotion Model of Pender could be applicable as a theoretical framework to identify major determinants of adherence to hypertension control recommendations.

**Figure 1.**
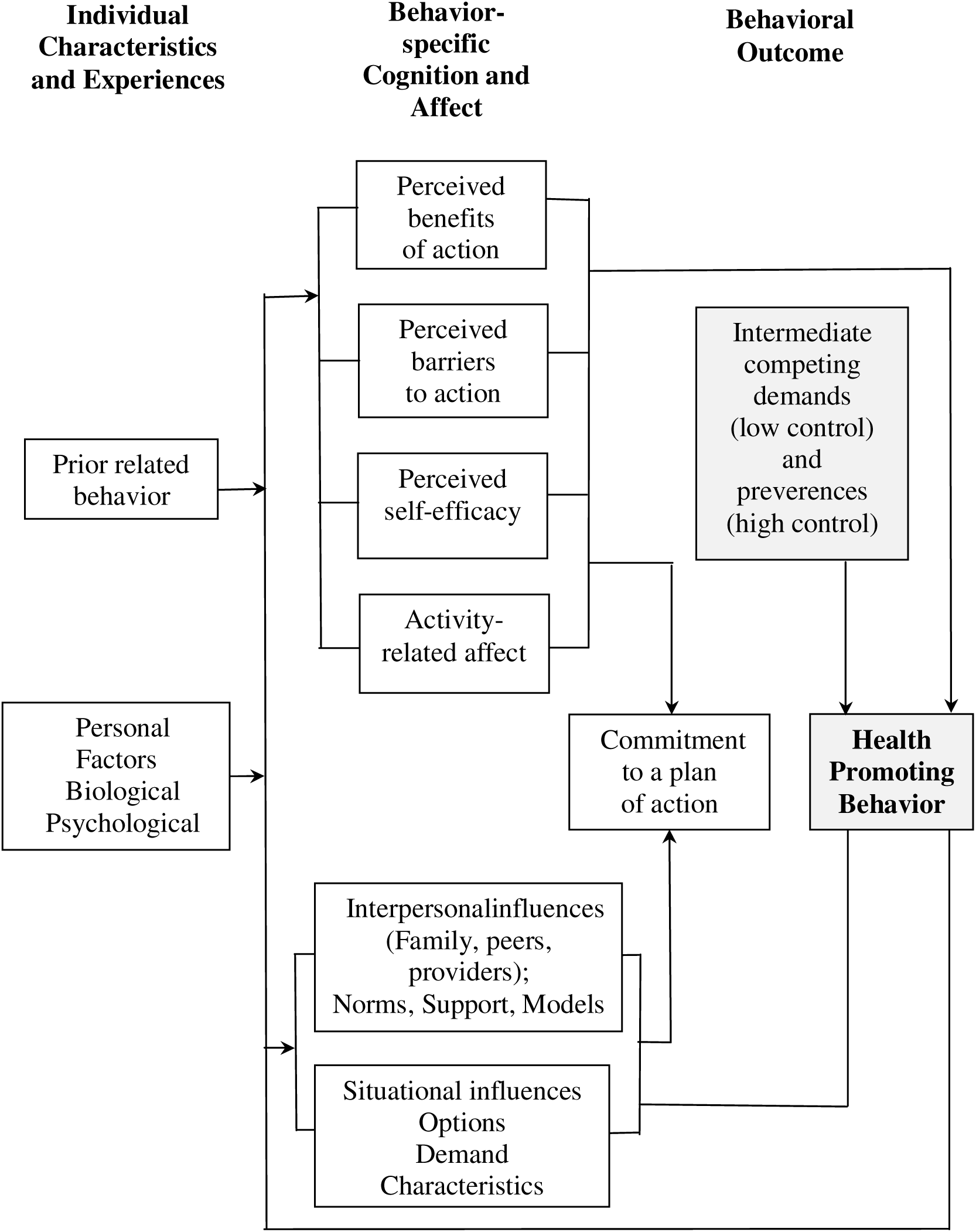
Shows the Health Promotion Model by Pender (1982)

#### Schematic Diagram

**Figure 2.**
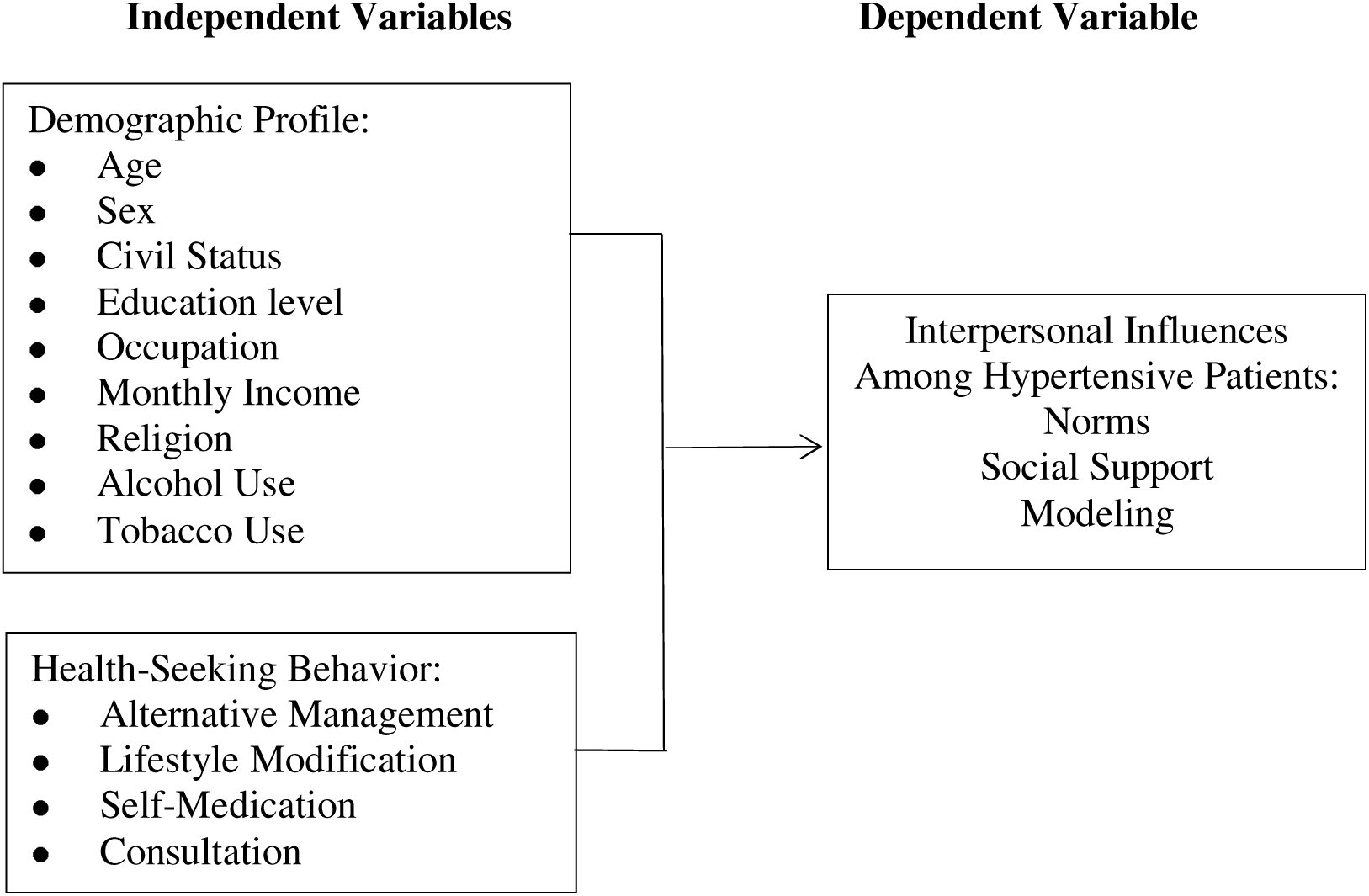
The Schematic Diagram of the Independent and Dependent Variables.

##### Statement of the Problem

The purpose of this study is to determine the health-seeking behavior and interpersonal influences among the hypertensive patients of Barangay Canitoan, Cagayan de Oro City. Specifically, this study aimed to answer the following questions:

1. What is the demographic profile of the participants in terms of:

1.1 Age,
1.2 Sex,
1.3 Civil Status,
1.4 Education level,
1.5 Occupation,
1.6 Monthly Income,
1.7 Religion,
1.8 Alcohol use, and
1.9 Tobacco use
2. What is the health-seeking behavior among hypertensive patients in terms of:

2.1 Alternative management,
2.2 Lifestyle modification,
2.3 Self-medication, and
2.4 Consultation
3. What are the interpersonal influences among hypertensive patients in terms of:

3.1 Norms,
3.2 Social Support, and
3.3 Modeling
4. Is there a significant relationship between the interpersonal influences, health-seeking behavior, and demographic profile among hypertensive patients?

##### Hypothesis

The investigators of the study hypothesized the following null hypotheses which will be tested at alpha 0.05 level of significance:

H_0_1-There is no significant relationship between the participants’ demographic profile and their interpersonal influences.

#### H_0_2- There is no significant relationship between the participants’ health seeking behavior and their interpersonal influences

##### Significance of the Study

Community. The community is considered an active partner rather than a passive care recipient. Therefore, this study will emphasize the community’s active role in managing individual health conditions and the interpersonal relationships that influence them throughout the course. Awareness of the deficits of their significant other’s role in their health will enable them to recognize areas that need improvement and be encouraged to adopt changes.

Nursing Learners. Immersing with the community will provide them with first-hand experiences that will contribute to an improved understanding of their client’s health situations, particularly how social contexts shape health-promoting behaviors. These learning will further aid them in rendering care in the community or healthcare setting.

Faculty. The faculty of the College of Nursing of Liceo de Cagayan University can benefit from this study as it discusses health-promoting behaviors that are influenced or result from interpersonal influences, thereby opening a different perspective and approach in the delivery of instruction to the students. Moreover, increased and positive awareness of their definite roles as nurse educators and how they influence other people can bring about change, not only in the academic environment but also in the community.

Future Investigators. Future investigators can use this study as a reference in determining the health-promoting behavior shaped by interpersonal influences, be it in investigating other health-promoting behavior for a different health condition. They can also use the findings from this study to correlate it with other variables in Pender’s Health Promotion Model.

##### Scope and Delimitation of the Study

The study focused on the health-seeking behavior and interpersonal influences of patients diagnosed with hypertension, residing in Barangay Canitoan, Cagayan de Oro City. The interpersonal influences involve norms, social support, and modeling. Consequently, the investigation of health-seeking behavior (early or late behavior) for hypertension includes, but is not limited to, alternative management, lifestyle modification, self-medication, and consultation. The investigators only determined which among these behaviors are practiced by the participants or not, and have not further investigated into deeper details such as the duration and frequency of the said health practices. The significant others present during the data collection were also briefly interviewed for the validation of the primary participants’ responses. The study was implemented during the second semester of the academic year 2022-2023, from January 2023 to May 2023.

On the other hand, the investigators have not further investigated the participant’s medical history, hypertension status, and level of knowledge about their condition. The study only determined the health-promoting behavior resulting from the hypertensive patients’ interpersonal influences.

##### Definition of Terms

To acquire a better understanding of this study, the following terms are operationally defined.

Age- is described in the number of years how old the participants are.

Alcohol Use-describes the status of the participants’ usage of alcohol regardless of the frequency and duration of habit. Includes only whether the client consumes alcoholic drinks or not.

Alternative Management-refers to participants’ management of their hypertensive condition through the use of herbal medicine instead of commercial medicine, as well as non-pharmacological techniques such as physical activity, healthy diet, and relaxation techniques.

Civil Status-defines the participants of the study as being single, married, separated or widowed/widower.

Consultation-refers to the hypertensive patient’s routine visitation or consultation with a physician in a clinic or in the Barangay Health Center for check-up purposes.

Education level-describes the highest educational attainment of the participants, which includes elementary, high school, college, and post-graduate studies.

Health-Seeking Behavior- is any activity carried out by the hypertensive patients to manage their condition, which includes: Alternative management, lifestyle modification, self-medication, and consultation (Johnson et al., 2021).

Hypertension- is blood pressure that is higher than normal and a consistent, constant elevation of systolic pressure above 140 mmHg and/or diastolic pressure above 90 mmHg (Tan & Thakur, 2021).

Hypertensive patients-refers to a person diagnosed with high blood pressure by a healthcare worker or is currently taking anti-hypertensive medications.

Interpersonal Influences- are cognition concerning behaviors, beliefs, or attitudes of others. These influences include norms (expectations of significant others), social support (instrumental and emotional encouragement), and modeling (vicarious learning). Families, peers, and health care providers are primary sources of interpersonal influences.

Lifestyle modification-refers to the participants’ healthy behaviors in managing hypertension including involvement in physical activity, eating a healthy diet, and practice of stress management or relaxation techniques.

Modeling- a way of learning that allows individuals to learn from the experience of others through observing others engaged in a particular behavior. Modeling influences include family, friends, famous personalities or trends from social media, television, radio, and healthcare workers that encourage a hypertensive patient to adopt health-seeking behaviors.

Monthly Income-refers to the countable income received in one month. The study will refer to the monthly income bracket established by the Philippine Institute for Development Studies (PIDS) in 2018:

Poor: Below P10, 957
Low-income but not poor: P10, 957 - P21,914
Lower Middle: P21, 914 - P43,828
Middle: P43,828 - P76,669
Upper middle: P76,669 - P131, 483
Rich: P219, 140 and above

Norms- the belief or expectations that determine and regulate appropriate behavior within a culture, group, or society. It is the expectations or expected behavior in managing hypertension, given by significant others from one’s family such as a partner, children, and other close relatives.

Occupation-refers to the profession, regular work, job, or principal activity, describing whether the participants are employed, self-employed, homemaker/housewife, student, unemployed, retired, or unable to work.

Religion- a group of organized values, customs, and ways of life that frequently revolve around the belief. It describes the religious group which the participants are affiliated with. The study uses generalized classification which are: Roman Catholic, Other Christian Churches, and Islam.

Self-Medication- refers to the participant’s intake of prescribed anti-hypertensive drugs for maintenance or other selected supplements.

Sex-identifies the participant as male or female.

Social Support- the support one receives from friends, family, peers, coworkers, and others on a physical, instrumental, and emotional encouragement. It includes tangible (money and medication supply) and non-tangible support (words of encouragement) in adopting health-seeking behaviors to manage hypertension.

Tobacco Use-Describes the status of the participants’ usage of tobacco smoking regardless of the frequency and duration. Includes whether the client smokes or not.

## Chapter 2 REVIEW OF RELATED LITERATURE AND STUDIES

This chapter presents a recommence of reviewed related literature and studies that have bearing on the conceptual bases of the present study. In here, there are literature which support study and give additional information by which this chapter has included.

### Age

The age group classification used in this paper was according to the classification introduced by the Kirklees Council (n.d.), recommended for survey questionnaires. These age groups are: 18 to 24, 25 to 34, 35 to 44, 45 to 44, 45 to 54, 55 to 64, and 65 and over. This is the most commonly used age band for consultation with adults.

Care-seeking behaviors provided a stepping stone to a successful management of non-communicable diseases (Gabrani et al., 2021). In turn, it is influenced by various factors which are socioeconomic conditions, age, genderm financial means, the individual’s health and illness perception, type of illness, and accessible health services. Hearn et al. (2019) stated that the elderly are perceived to be dependent on their families or head of household. Adults and elderly who have chronic conditions received care from multiple providers in over different settings; however, this care is often poorly structured (Elliot, 2018).

A study conducted in Albania by Gabrani et al. (2021) has revealed that the prevailing chronic conditions among adults and elderly were high blood pressure, which is 44% in adults and 74% in the elderly. Older participants of the study were more likely to have a low education level, which is primary education or below, compared to younger participants. Also, the coverage of health insurance was higher among them. Furthermore, those in the adult age group who participated in the study were more likely to frequent a hospital which comprised 46%. On the other hand, those in the elderly group were more likely to seek governmental PHC health centers. In terms of the frequency of health-seeking behavior, services utilization was higher among the elderly (52%), who were predominant in seeking care one per month, while adults (46%) only sought care several times per year for the control of their condition. It was revealed that the presence of self-medication and the belief that the health problem would resolve without medical treatment were the main reasons for not seeking care.

Community-based health promotion activities have multiple benefits to improve physical functions, spiritual satisfaction, and a sense of accomplishment among older adults (Liljas et al., 2017). A cross-sectional study conducted by Chen and Hsieh (2021) investigated the health promotion activity among older adults in Taoyuan City. The findings indicated that age, perceived benefits, and self-efficacy were significant predictors of participation in health-promotion activities. Majority of the elderly people who participated in the said study had chronic diseases, comprising 76.3%. Age and self-efficacy had relatively higher contributions toward participation in health promotion activities.

The importance of social capital in improving the health and well-being of many social categories was now established as a social construct that has undergone extensive research. In Quezon, Nueva Ecija, in the Philippines, 120 people who were 60 years of age or older were randomly chosen to participate in a survey. The study examined the senior population’s social capital in a rural setting and how it related to their health condition and health-seeking habits. The following five domains, general trust, social support, social networks, social involvement, and close ties were used to measure the social capital of elderly people. The elderly defined themselves as most healthy when they can undertake everyday activities, whereas they described themselves as least healthy when they have experienced chronic conditions in the past three months, according to results from correlation and multiple regression analyses. Their top health-seeking behavior was self-care, which included eating well, exercising, and having fun, but it should be mentioned that elderly people preferred to practice better self-care if they have strong relationships and participate in social activities. Despite the fact that elderly people rarely seek medical attention, the findings suggested that as social support grows, so does the frequency of the aforementioned health-seeking behavior (Bernardo & Tolentino, 2019).

In terms of physical activity, the study of Rababa et al. (2021) has yielded low mean scores of physical activity among older adults, indicating that they were not regularly engaged in physical activity. A decline in physical activity often resulted from age-related musculoskeletal changes such as decreased muscle and bone mass and reduced muscle strength (Fanning et al., 2020).

To encourage positive health behavior change and to prevent the beginning of more impairment, morbidity, and even premature mortality in the future, future health behavior interventions must concentrate on the elderly and people with physical disabilities. To better understand how health behavior changes, future studies should look at the effects of individual characteristics (such as personality, social network structure, demographics, health beliefs, and comorbidity) in addition to age and health status as predictors of different health behavior trajectories (Zanjan et al., 2020).

### Sex

It was revealed that gender was significantly and positively correlated with health. Men had better health than women, whether it is subjective health evaluation or objective health. A possible explanation was given, and that is, when comparing men with women, the latter devoted more time and energy caring for their children and doing household work. Moreover, they were expected to handle job affairs very well. In turn, women encounter difficulties in balancing family responsibility and career development, affecting both subjective and physical health status (Long et al., 2020).

On the other hand, men were assumed to be more at risk for cardiovascular disease from hypertension than women. Studies have revealed that women have a greater overall incidence of hypertension-related cardiovascular disorders than males, and this was particularly true during the menopause, when the prevalence of hypertension and its complications rises suddenly and sharply. Additionally, it was noted that different anti-hypertensive groups had varied effects on men and women (Marijana et al., 2019).

According to Tadic et al. (2020), Pre-hypertension prevalence was 46.4%, and hypertension prevalence was 37.3%. Also, 49.7% of hypertensive individuals were aware of their condition, and 71.5% were taking antihypertensive medications on a doctor’s prescription. Furthermore, 38.9% of those who received treatment had their blood pressure under control. In the revised model, increased awareness was positively correlated with older age, female sex, and history of diabetes mellitus. Only high levels of physical activity, smoking, and diabetes can predict treated high blood pressure. Higher education, female sex, and younger age were all associated with managed hypertension. The awareness and management of hypertension were substantially connected with health insurance.

While recent health reforms in the Philippines have made an effort to lessen the financial burden at the point of care for people living in poverty, research showed that women still relied on their social networks for financial support when health insurance coverage was insufficient to cover the direct and indirect costs associated with the pathway to healthcare. Additionally, these results illustrated how crucial social networks are in this situation for easing the financial and social difficulties related to women’s access to health care services at various periods of their lives. These findings implied that fostering and enhancing social networks among poor women can boost ongoing efforts to put UHC in the Philippines (Luu et al., 2022).

In the entire world, cardiovascular diseases (CVD) continued to be the largest cause of death. According to studies, men experience symptoms ten years earlier than women do, and they are less likely to adopt healthy living habits than women are (Botabara-Yap et al., 2021). The body of knowledge about men’s health-seeking behavior in academia has been properly expanded by this study. It went into great detail about a variety of elements that affect men’s pursuit of health. With a focus on men in a private higher education institution in Nigeria, it explicitly examined if educational attainment has an impact on attitudes toward seeking health. The study has demonstrated that health-seeking behavior is a complex junction of personal health perceptions and cultural and societal norms that are widely followed in all civilizations. Men’s health-seeking behavior was greatly influenced by their masculinity, cultural beliefs, social conventions, and economical variables (Olanrewaju et al., 2019).

Age, female gender, fast food consumption, and animal fat intake are linked to missed targets for blood pressure control and hypertension-related hazards. Males who frequently drank and smoked also tend to have poor blood pressure regulation. Regular consumption of fruits and vegetables was linked to improved blood pressure management and a lower risk of overall hypertension. The observed unsatisfactory BP control despite pharmaceutical adherence pointed to the need for lifestyle changes in addition to antihypertensive drugs. By combining lifestyle change, medication adherence, and expert-tailored information delivery, interventions should target modifiable risk factors that are exacerbated by aging and unhealthy lifestyles (Kimani et al., 2019).

Male adolescents with extreme obesity who sought therapy had more negative metabolic and behavioral risk factors than female adolescents. Findings suggested that both genders should have their lifestyle behavioral markers extensively evaluated, and that potential gender-related differences in risk profiles should be taken into consideration in future treatment plans (Barstad et al., 2018). A study by Reckelhoff (2018) proposed that men and women receive similar therapy; however, this was not the situation in reality. Women were more conscious of hypertension than men were, and women were more likely to have it until beyond menopause. Recent research has examined the roles played by the kidneys, the renin-angiotensin system, relaxin, and developmental programming in the mechanisms underlying sex/gender differences in hypertension.

Gender disparities in risk factors, awareness of, treatment for, and control of hypertension were widely documented. Men and women had distinct variances in the epidemiology and clinical features of hypertension. While the control rate for men and women on antihypertensive medications were similar, gender differences have been linked to the prevalence and causes of hypertension and prehypertension (Song et al., 2019).

### Civil Status

The classification of civil status used in this paper was according to the one presented by the Philippine Statistics Authority (2017), which are: single, married, separated, and widowed/widower. One of the important social factors associated with human health and longevity is marital status. A study conducted by Kim et al. (2018) investigated the relationships between marital status and health behaviors and illness among Koreans who are middle-aged, and tested whether gender differences existed in the said relationships. First, the health behaviors of the participants were collected which included the following: physical activity, smoking status, alcohol consumption, sleep duration, and participation in health examinations. In terms of nutritional intake, the results revealed that total energy intake was lower in men and women living without compared to those who lived with their spouses. Moreover, higher rates of smoking and binge drinking, worse sleep duration, and non-participation in regular health examinations were noted in both sees living without their spouses. Women of the same category showed higher rates of diseases such as hypertension, diabetes, hypercholesterolemia, hypertriglyceridemia, depressive mood and suicidal ideation.

Pandey et al. (2019) indicated that healthcare utilization patterns were influenced by marital status. Health care costs among beneficiaries of medicare are half compared to those who are not married. Also, socio-demographic factors served as an important determinant of health care utilization and unmarried individuals have reduced access to resources that may affect the latter. Lastly, previous research suggested that being married is predictive of better health status. The findings of the study asserted that marital status is an important predictor of healthcare utilization. This is because spouses can function as caretakers at home, which excluded the need for formal healthcare. Moreover, they can assist with medication adherence, in the preparation of healthy meals, and ensuring attendance in appointments with physicians.

Moreover, a long-term follow-up study was conducted by Ramezankhani et al. (2019) to determine the associations of marital status with diabetes, hypertension, cardiovascular disease and all-cause mortality. The findings revealed that being never married in men was associated with higher risk of hypertension and all-cause mortality. On the other hand, when comparing women with married status, those who were widowed had an association with a lower risk of type 2 diabetes mellitus. Also, never married women had a lower risk of hypertension.

On another note, the lack of physical activity posed substantial consequences for health and mortality risk (Aggio et al., 2020). The relation of marital quality to physical activity among men and women were investigated by Thomas et al. (2022). The general finding of the study indicated that marital support and strain were related to higher chances of more frequent active exercise and walking. It became increasingly significant to higher levels of walking frequency among men. They supported that spouses have the potential to be powerful sources of influence that serve as influential factors as individuals age.

According to Segawa et al. (2021), the prevalence of hypertension was higher in married females or those who were separated, divorced, or widowed compared to females living together with their spouses. Also, the prevalence of the said disease differed in terms of marital status and sex. The study of Son et al. (2022) revealed divorce as the marital status that strongly affects hypertension. It had a strong association with high blood pressure in males and a higher risk in females. Defianna et al. (2021) also argued in their study that females were economically highly dependent on male, and being divorced as well as poor were identified as risk factors.

### Education

The classification of educational attainment used in this paper was based on the education system in the Philippines presented in the Scholaro Database (n.d.), which are: Elementary (Primary level), High school (Secondary level), Vocational, College (Undergraduate level), and post-graduate level.

Giena et al. (2018) claimed that education has a positive correlation with health-promoting behavior. They explained that participants with high education levels were likely aware of their health behavior. This is due to the result of their study that older adults with high education are willing to seek information regarding their condition.

Moreover, understanding information about hypertension and healthy behaviors becomes easy.

The level of development including income, level of education, accessibility and transport communication, mass media exposure, age, se, quality of health care system and the availability of services directly or indirectly influence human behavior and attitudes toward health care (Lalmalsawmzanva & Lalrohlua, 2018). The reviewed literature shows that health seeking behavior among women in many parts of the world is affected by low literacy, lack of awareness, and low status.

The level of education of the participants in the study of Adongo (2018) has shown direct influence of the participants’ understanding of good health behaviors. This means that the more educated people are, the more they understand their health conditions and therefore seek solutions. It further indicated that educated people knew the importance and benefits of engagement in health promotion behaviors, and they had better access to different health promotion resources. Thorpe et al. (2019) also stated that educated people were more aware of the negative consequences of unhealthy lifestyle and practices on their health.

A study conducted by Long et al. (2020) analyzed the relationship between education and health status of Chinese residents. The study found that education promoted the subjective and objective health of the residents. Moreover, education has the capacity to promote health through improved mental health, economic status, and healthy behaviors. The investigation further revealed that the higher the level of education was, the higher the participants’ self-reported health, and the frequency of health effects were lower.

Mediums for health education were also investigated by several studies. It has been noted that educational reinforcement through the use of technological tools, such as mobile applications, was a facilitator of diabetes self-management and could be used as an intervention to close gaps in diabetes self-management. To further improve diabetes self-management, it is crucial to develop new strategies that address the financial burden, work- and environment-related variables, as well as diabetes misery (Adu et al., 2019). The most important influencing elements were education level and diabetes education programs. According to the current study, regular structured training sessions on diabetes and its consequences were necessary to help patients lead fulfilling lives (Rahaman et al., 2017). It was also found that not all health education methods have a positive correlation to health-care seeking behavior. The methods that showed positive correlation were lectures, public consultation, and online education. Consequently, publicity material and bulletin boards showed no correlation. It explains that residents are more inclined to face-to-face learning (Li et al., 2020).

### Occupation

The classification of occupational status used in this paper was based on the one stated by DeFranzo (n.d) in Snap Surveys, which are: Employed, self-employed, homemaker, student, unemployed, retired, and unable to work.

The study of Barbini et al. (2017) examined the professional risk factor for hypertension, particularly with regard to aging of the workforce. The participants were workers for high-speed trains in the Tuscany region. The analysis of data showed that drivers of heavy vehicles had the highest prevalence of hypertension. Moreover, the main occupational risk factor associated with hypertension was a job that requires prolonged attention. The findings also revealed that hypertension was detected in more than half carpenters or welders and managers. Other characteristics also showed a significant risk factor, such as aging, obesity, irregular sleep, irregular eating, and night shifts.

The work people do affects their risk of hypertension, and it was an important social determinant of the said disease. Chantarat et al. (2022) cited three occupational determinants of hypertension; these are: job insecurity, job loss, and the psychosocial work environment. Acevedo et al. (2019) supported that workers who have experienced job insecurity or job loss are more likely to have hypertension than those who have not. However, the degree as to how these factors affect blood pressure varies according to a worker’s gender, age, the time of job loss, and how long the worker has been unemployed.

Qin et al. (2022) studied the association of socioeconomic status with hypertension prevalence and control in Nanjing. The results yielded that there was a high prevalence among subjects who were unemployed and retired. Moreover, higher odds of hypertension were observed in the mental laborers or students. It is further argued that hypertensive factors such as social psychological factors, working overtime, exposure to high temperature, and noisy environment are common among manual laborers; thus, rendering them a higher prevalence rate of hypertension.

In terms of health seeking behavior, Latunji and Akinyemi (2018) studied the factors that influence it among civil servants in Ibadan, Nigeria. Two groups of people were identified; the first group were those who sought formal health-care services, and the second group were those who focused on illness response or health-seeking behavior. The findings stated that appropriate health-seeking behavior was high among civil servants. Those who belonged in the poorest quartile were six times more likely to have inappropriate HSB than those who belonged in the richest quartile. They suggested that policy should be formulated to improve health-seeking behavior among cadre workers and those with lower levels of education.

Occupational status and working conditions were also investigated by Qi et al. (2020), which showed the relationship between occupational status and health inequality among workers in China. The results of the study yielded that low occupational status was associated with poorer self-assessed health, and an increased incidence of injuries related to work. Moreover, those who have high occupational status frequently reported more occupational diseases. The authors also argued that the longer the length of education, the likelihood of work-related injury is lower.

The study of Adei et al. (2022) compared the health-seeking behavior between formal and informal sector workers in the Kumasi metropolis of Ghana. The results of the study revealed that 33.5% of the participants were practicing good health seeking behavior, which involves seeking for healthcare in a health facility. They further analyzed that most of the informal sector workers sought healthcare from pharmacies, herbalists, spiritualists, first aid at the workplace, and home treatment when experiencing occupational disease. Those who work in the informal sector who spent more than an hour at a health facility were found to have greater chances of adopting health-seeking behavior compared to those who spent less than an hour.

### Monthly Income

The Philippine government utilizes the income class bracket established by the Philippine Institute for Development Studies (PIDS) in 2018 to categorize families into social classes. According to Celia Reyes, the president of PIDS, the pooled monthly income of families is what identifies them as rich or poor. The following are the identified social classes based on income brackets: Poor: Below P10,957 monthly income; Low-income but not poor: P10,957 to P21,914 monthly income; Lower middle: P21,914 to P43,828 monthly income; Middle: P43,828 to P76,66 monthly income; Upper middle: P76,669 to P131,484 monthly income; Upper middle but not rich: P131,483 to P219,140 monthly income; and, Rich: P219,140 and above monthly income. As of 2018, statistics from the PIDS revealed that 8.4 million families belong to the low-income bracket, and 7.5 million belong to the lower-middle income. On the other hand, 4.3 million families are categorized to the middle and upper middle class. Upper but not rich class comprised 358, 000 families, while 143,000 families were classified as rich.

As stated by Qi et al. (2022) in his study among adults in Nanjing, China, socioeconomic status has an important role in hypertension prevalence and hypertension control. It was suggested to focus on people belonging to the vulnerable lower SES groups in terms of strategies for prevention and control. The odds regarding controlled hypertension were decreased among adults with lower annual household income when compared to those who have higher income. Furthermore, people with higher household income were more likely to have a healthy environment in terms of living or work; thus, enabling a higher chance of a delayed onset of developing high blood pressure or adequate control of high blood pressure. Musinguzi et al. (2018) also supported that patients with greater economic resources are likely deemed to seek treatment from doctors in private facilities and spend a more considerate amount, compared to those belonging to a low socioeconomic index.

The study of Hogg (2022) looked at how hypertensive people perceived their treatment adherence, attitudes toward hypertension, and understanding of the condition. According to the study, health interventions posed a slight challenge with impoverished populations as compared to the general population. Moreover, when an individual believes their hypertension diagnosis and that the potential complications of the disease are serious and believe that the treatment is effective, they are likely to adhere to their treatment if the perceived barriers to treatment are only minimal. On the down side, impoverished populations might face more perceived barriers than general populations. Someone facing impoverishment is likely to use their financial resources to meet basic needs such as food, water, and clothing before purchasing medication, particularly when they have little understanding of the importance of the medication and complications of hypertension.

Rababa et al. (2021) also found significant differences between participants based on their family income in terms of nutrition and exercise behaviors. It was indicated that older adults with higher income had higher levels of nutrition and exercise. Consequently, older adults with low socioeconomic status received less information about HPB, making them less engaged with it (Bernardelli et al., 2020). The burden of hypertension management is further supported by Zhou et al. (2017). It was stated that raised blood pressure has transitioned from a significant burden in high-income countries to a concern that is now highly prominent among low- and middle-income countries over the past three decades. Allen et al. (2017) argued that people in the low socioeconomic groups were more likely to consume unhealthy diets, use tobacco and alcohol, and were more likely exposed to air pollution. Upon the diagnosis of hypertension, those who belonged in low socioeconomic status were less likely to afford out-of-pocket expenses to purchase antihypertensive medication, which eventually led to uncontrolled hypertension (Schutte et al., 2021).

### Religion

The classification of religions used in this paper was according to the top three religious groups in the Philippines presented by Northern Illinois University (n.d.), which are: Roman catholic, other Christian denominations (non-catholic groups), and Islam.

Religious people such as clerics and other religious entities can be a good source of support for women in the community (Morowatisharifabad et al., 2019). Investigating the attitudes and health perceptions of faith healers in Ghana’s Kumasi Metropolis was the main goal of the study. This is essential because despite its importance, faith healing was rarely examined in studies on healthcare. In an era where bio-western medicine was presented as the cure for the majority of diseases, understanding how and why people sought alternative treatments, in particular faith healing methods, may assist to build more effective health care interventions (Peprah et al., 2018).

The study of Heinert et al. (2019) assessed feedback on a church-based hypertension intervention. The results yielded that the most common facilitators for hypertension control were social support, knowledge on how to control hypertension, and resources from the community. On the other hand, the common barriers were lack of knowledge regarding hypertension, negative experiences with primary care providers, and lack of awareness of the disease. On another note, Ogedegbe (2018) indicated that implementation of a lifestyle program in a faith-based setting was found to be effective in improving blood pressure control and can possibly be a routine practice in places where worship is done.

Religiosity posed benefits on the attitudes, motivations, goals, social interaction, and perceptions of people regarding wellness which was associated with attendance in church (Bruce et al., 2017). Moreover, it was suggested that those who were deeply involved with their religion were likely to have health outcomes, considering that their lifestyle is in line with their religious belief. Davis (2021) revealed in his study that religious people had higher health-awareness, which prompted them to adopt actions such as staying active, managing stress, getting, resting, and choosing healthier food. The identification of health awareness was identified through prayer, Bible, and social support. The participants of the study claimed that the Bible has provided them with instructions on how to take care of their body and adopt a healthy diet. Social support, including church services, has provided participants with advice on hypertension self-care, accountability and support, health education, and message as to how God wanted them to care for themselves.

### Alcohol Use and Tobacco Use

According to Nagao (2021), alcohol and smoking were seen in a combined perspective as both result in a transient increase in blood pressure and onset of persistent hypertension. They further indicated that the increase in the amount of smoking also contributed to an increase in the risk of developing hypertension. If an individual can be motivated to moderate alcohol consumption and quit smoking, more optimal health guidance can be given. The study by Yadav et al. (2021) concluded that regardless of wealth, quintile, residency, and dietary preferences, alcohol, and smoking lead to greater chances of being hypertensive. The proportion of hypertensive participants among alcoholics or smokers who are diabetic was higher than non-diabetic alcoholics or smokers. The study also stated that obese individuals tend to have higher chances of getting hypertension notwithstanding their addiction status. This is because obesity is a major contributor to raised blood pressure.

Type 2 diabetes is 30% to 44% more likely to occur in smokers than in non-smokers, and the risk increases with cigarette consumption. Numerous biological mechanisms, such as smoking’s effects on cortisol levels, central obesity, inflammatory markers, oxidative stress, insulin resistance, and an increase in fasting blood glucose, have been proposed as biological explanations for the causal relationship between smoking and the onset of type 2 diabetes (Bush et al., 2017).

The most popular way to consume tobacco is by smoking, primarily in the form of cigarettes and burnt tobacco. Even if smoking rates are falling in some nations, public health is still seriously threatened by it, especially in eastern Europe and central and south-east Asia, which have the highest rates of smoking worldwide. By 2050, the World Health Organization (WHO) predicts that there will be 1.5 billion smokers worldwide. Every organ and system in the human body is affected negatively by smoking cigarettes, which is widely recognized to cause a wide range of illnesses and problems (Campagna et al., 2019).

Smoking raises the risk of T2D for active smokers by 30%–40% compared to nonsmokers, suggesting that smoking cessation should be highlighted as a crucial public health strategy to battle the global diabetes epidemic. The World Health Organization supports quitting or avoiding smoking as part of their lifestyle recommendations since it identifies smoking as a preventable risk factor for T2D (Maddatu et al., 2017).

The rates of abstinence and/or the discontinuation of heavy drinking have significantly increased in studies with an average duration of 6 months, according to thorough meta-analyses of randomized controlled trials using FDA-approved drugs to treat AUD. It is important to recognize that all participants in those clinical trials received either protocol-specific behavioral treatments or the nonpharmacological therapy for AUD that is typically offered in a particular context. As a result, the drug showed a noticeable advantage over the placebo in terms of drinking results (Mason et al., 2021).

If beer’s potential health advantages are taken into account, its alcohol concentration is a legitimately important matter. In more economically developed nations, alcohol is the primary cause of liver illness and is linked to high levels of social dysfunction, decreased productivity, and bad health (Mellor et al., 2020).

### Health-Seeking Behavior

Health-seeking behavior is part of the broader concept of health behavior, which includes activities undertaken to maintain good health, prevent disease, and manage any deviations from good health. In the study of Latunji (2018), there were two groups that described the factors that influence health seeking behavior during the time of illness. The first group is made up of studies that focused on the use of the formal system or on health-seeking behavior. Studies that fall into this category involved the development of models that described the series of steps people take toward health care. These models were sometimes referred to as “Journey models.”

Although there were several variations of these models, Andersen’s Health Belief Model and Health Behavior Model were often used as the basis in discussions of BHS. The second group included studies that focused on the disease response process or health-seeking behavior. These studies showed that the decision to engage in a particular medical channel is influenced by a variety of factors such as socio-economic status, gender, age, social status, type of disease, access to services and the perceived quality of services. The majority of studies in this second category focus on certain types of determinants that stand between patients and services, such as geographic, social, economic, cultural, and organizational factors. For example, access to healthcare facilities, socioeconomic status, and perceived quality of services was found to have been important factors influencing care-seeking decisions in different segments of the population.

Musinguzi et al. (2018) claimed that there was a delay in seeking medical care for hypertension, and was only often sought when complications of the disease occur such as stroke, heart attack, heart failure, and kidney failure. Thus, this signifies a need for reinforcement of health-seeking behavior. However, minimal health-seeking behavior was observed even among those who were symptomatic and patients who were aware of their condition, resulting in low-controlled hypertension. Some of the options made by patients in managing hypertension include visiting a public or private health facility, self-medication, used home remedies, and these decisions were dependent on factors that were personal, experiential, and sociocultural (Gupta et al. , 2020).

A study conducted by Jabar (2018) investigated the factors that influenced health-seeking behavior among overseas Filipino workers. He concluded that the variables that predict health-seeking behavior included the perceived health status, history of sickness for the past two years, education level, age, and number of children. The income and sickness were significant predictors of seeking care from a traditional health provider. Consequently, sickness in the past two years, sex, perceived health status and age are what influences an individual to resort to self-medication. On the other hand, the aforementioned variables, but including income, are what predicts a visit to the hospital. The author summarized that health promotion should consider the variety of socio-demographic and personal characteristics of individuals in order to increase health seeking.

The study led by Phromjuang (2021) focused on the health-promoting behaviors of elderly hypertensive patients and to identify factors that influenced health-promoting behaviors in elderly hypertensive patients. Subjects were elderly hypertensive patients attending a local health center affiliated with Watbot Hospital. Data were collected through investigator interviews and questionnaires. Seven questionnaires were included, including demographic information, social support, interpersonal influence, self-esteem, health beliefs, hypertension knowledge, and health-promoting behaviors. A hierarchical stepwise multiple regression analysis was performed.

In general, there were two factors that explain differences in health-promoting behaviors of older people with higher levels of hypertension. Two factors that predicted health-promoting behavior were interpersonal influence and social support. The study suggested that nurses or critical caregivers should promote health-promoting behaviors in the elderly by providing them with love and care. When older adults are praised or have a good role model for health promotion, they are better able to control their blood pressure. Examples of health-promoting behaviors include the use of medical services, compliance with medical regimens and self-directed health practices (Giena et al., 2018).

### Alternative Management

According to Musinguzi et al. (2018), most patients who were seeking complementary medicines used herbs, reporting that they have consulted traditional healers for their hypertensive condition. Patients have considered it as readily accessible and comparably affordable. Neamsuvan et al. (2018) stated that hypertension is the most important risk factor for developing cardiovascular, renal, and ocular disease in Thailand, morbidity and hospitalization due to hypertension are increasing in the modern public health system. However, some Thais have come to use herbal medicines for health management. The study provided knowledge about the treatment of hypertension, and examined the use of plants in the treatment of high blood pressure by folk medicine practitioners. Most healers believed that high blood pressure was caused by a disruption of the fire and air elements in the body. Pungent, mildly flavored medicinal plants were applied that might treat air element problems. A total of 62 species are used to treat high blood pressure. Most of the plants belonged to the family Asteraceae, Piperaceae, Rutaceae, or Zingiberaceae (4 species each). Herbal medicines were preferably prepared by cooking (78%) and consumed by drinking one cup of tea before three meals daily (26%). The identification of medicinal plants tested by experienced folk medicine physicians provides an opportunity for people to select and consume local herbs that are readily available in their area.

The study of Rondilla (2021) discussed the lived experiences of health seekers acquiring folk medicine services in Quiapo and described the significance behind their acquisition of these services. Local studies showed that adherents of folk medicine practices in the Philippines are mostly married Catholic women of low economic status and low educational attainment. Similar to foreign studies have also shown that consumers of traditional and complementary medicines are often women and married, whose age is in the average range of fifty years and whose level of education has a high school diploma. These sociodemographic profiles were all evident in the findings of the present study. Another study also found that rural populations had unique health-seeking behaviors and had conflicting views on medical pluralism, which explains the behavior of these health-seeking individuals.

The availability of medicinal plants in Quiapo and how Filipinos protected them state a lot about the country’s greater cultural diversity. Additionally, for centuries, most cultures had used herbal medicines as effective therapies to treat and even prevent various diseases and conditions. With the modern advancements of the internet, customers now enjoyed wider access to herbal products around the world. The influx of consumers using Quiapo folk medicine services confirms the fact that a large part of the population still uses herbal medicine. One of the main reasons people used traditional medicine in previous studies is that it is used in addition to conventional medicine. Its high popularity and ease of access are the main factors that lead to self-medication. People who self-medicate seemed to rely heavily on natural and alternative medicine compared to their pharmaceutical counterparts. They assumed that since these were considered natural remedies, this automatically equates to safety. It is disturbing how alternative medicines were more readily available and easier to purchase without the need for a legitimate prescription. The resurgence of public interest in folk medicine has been attributed to various claims about the efficacy of plant or herbal medicines. The trend is currently towards the “natural”. It is likely that people’s growing perception of herbaceous plants has led to their use as both a curative and a preventative.

Another study by Liwa et al. (2017) investigated Tanzanian adults admitted with conditions related to hypertension. It is found that there is a frequent use of herbal and other alternative medicines, and many of these adults were using herbs specifically for hypertension. In this study, nearly 70% of adults reported having visited a traditional healer, and more than 75% said they had used herbal remedies in the past. A quarter of all adults currently use herbs specifically to control their hypertension. The use of herbal and alternative medicine by African inpatients has never been described in detail before, according to this study. Findings from hypertension clinics in South and West Africa as well as community-based studies in Eastern and Western African nations show a pattern that is generally similar to this one. A quarter of African adults with hypertension reported using traditional medicines in each of these settings, the majority of whom were herbal remedies. Collaboration with traditional healers should be looked into as a potential path for enhancing hypertension care because herbal medicines in particular are such a significant component of hypertension self-management throughout Africa.

The study of Adeniyi et al. (2021) stated that Complementary and alternative medicine (CAM) has been practiced for centuries all over the world and includes a wide range of treatments like dietary supplements, herbal remedies, traditional Chinese medicine, acupuncture, mind-body medicine, and therapeutic massage. A survey that examined the use of herbal remedies among rural and urban Jamaicans of varying socioeconomic groups found that 100 percent of the participants used herbs. The survey results and the themes from the focus group discussions (FGD) shed new light on participant perceptions of their propensity to use CAM and offered insightful new information.

Palileo-Villanueva et al. (2022) also argued that there are many different types of conditions that were treated with traditional, complementary, and alternative medicine (TCAM). It is especially prevalent in low-income individuals in low- and middle-income countries (LMICs) economic and social standing. The study examined the prevalence and characteristics of TCAM used for hypertension, its determinants, and its association with hypertension management outcomes and wellbeing among low-income adults in two Southeast Asian countries at different levels of economic and health system development in order to better understand the patterns and impact of TCAM used on the management of noncommunicable diseases in the populations of the Philippine Islands and Malaysia. The management of hypertension is carried out by TCAM, a small but not insignificant portion of afflicted residents of low-income neighborhoods in the Philippines and Malaysia. Still, there is proof of the effectiveness and safety of TCAMs that are frequently used. To make sure they are used, information on modalities and on their possible interactions with traditional medications is required to the best advantage. In the interim, national health systems must continue to work to increased access to primary care services, and healthcare professionals seeking to effectively treat hypertension from a patient-centered perspective must do the same acknowledge that their patients had been receiving advice treatment that could influence from a variety of sources their awareness, comprehension, and ultimately the control over their condition.

### Lifestyle Modification

The foundation of preventive management in individuals diagnosed with hypertension is lifestyle modification (Buda et al., 2017). Evidence from the literature of Sanchez et al. (2019) showed the benefit of lifestyle and antihypertensive medication therapy to lower blood pressure. However, the study of Gupta et al. (2020) yielded in their findings that many patients only had limited knowledge and certain misconceptions about the necessary lifestyle modification to lower BP. This could be explained when only few patients from their study identify the lack of physical activity and obesity as risk factors. The patients stated that they felt rest is needed more and physical activity should be avoided. Since hypertension is asymptomatic, it may lead some people to avoid seeking healthcare not until their condition eventually interferes with social or personal activities such as work and maintaining household works (Satish et al., 2019).

Hypertension, being chronic in nature, necessitates patients and their respective families to participate actively in their self-care, which involves activities like taking medication, becoming aware of complications of the disease and danger signs, having follow-up regularly, and most importantly, making lifestyle changes and managing changes in emotion (Gupta et al., 2020). Buda et al. (2017) studied the lifestyle modification practices and associated factors among patients diagnosed with hypertension in South Ethiopia. The findings revealed that only 27.3% percent of the participants practiced the recommended lifestyle modification or non-pharmacological approaches, which includes weight reduction, salt restriction, and physical activity, cessation from smoking, and alcohol abstinence. The suggested BMI for an adult (over 18 years of age) is less than or equal to 18.5-24.9. Additional information also indicated that those who had basic knowledge about hypertension, in treatment for 5 to 10 years, had a family history, and informed by a health professional practiced good lifestyle modification. It was concluded that patients should be educated on the recommended lifestyle modifications to be able to help them control blood pressure.

Adequate physical activity has been proven to have various health-promoting effects, and has also a direct effect in reducing blood pressure (Buda et al., 2017). Light exercises and other exercises such as brisk walking, jogging, cycling, and swimming can be done to help lower or control high blood pressure (Triangto, 2021). It is further explained that the time-period involved does not merely define a good exercise, but rather the duration and performance of the exercise in a related manner, not being rushed. The World Health Organization (WHO) recommends 30 minutes of exercise a day, which totals to 150 minutes per week.

Ma et al. (2021) studied physical activity and reduced dietary sodium intake for preventing hypertension and chronic disease among Filipino-Americans. It was emphasized that together with physical activity, dietary factors show a central role in hypertension development. Dietary salt intake has been associated with hypertension for a long time now. However, there are no studies yet that quantified the level of salt intake among Filipino Americans. Nevertheless, Asian Americans had an increased salt sensitivity experience. Grillo et al. (2019) also supported that not only reduction in dietary sodium lowers blood pressure, but also decreases the incidence of morbidity and mortality from cardiovascular diseases. Irrespective of sex and ethnic group, a prolonged reduction of salt intake caused a fall in blood pressure among hypertensive and normotensive individuals. This is due to the fact that high sodium intake is related to retention of water in the body and increased peripheral resistance.

Lorenzo and Evangelista (2021) studied the psychosocial correlation of hypertension among Filipinos who are primarily living in rural areas. The study introduces self reports of depression, anxiety, stress, and poor health-related life quality have been linked to an increased risk of hypertension (HTN). The study reveals that HTN was correlated with higher depression and lower physical-health related quality of life. Stress was the only source of hypertension for men, and both lower physical and mental health related quality for women. Lee et al. (2020) studied the effect and acceptability of Mindfulness-Based Stress Reduction Program on patients with hypertension. The mindfulness-based stress reduction program (MBSR) is the commonly used mindfulness-based interventions (MBIs), which trains a participant to develop awareness of paying attention in the present moment with self-understanding, wisdom, and compassion. The results yielded in the post intervention that MBSR decreased systolic and diastolic pressure by 6.64 mmHg, 3 to 6 months following the recruitment.

Lastly, A large-scale study was conducted by Gordon and Mendes (2021) to examine the effects of stress and emotion in blood pressure in daily life, utilizing a digital platform. It was found that older adults showed greater consistency between self-reported stress and physical responses compared to younger adults. Blood Pressure increased with negative emotions that cause high arousal such as anger, and decreased with positive emotions that are low arousal such as contentment. To summarize, higher demands or “stress” causes an elevation in blood pressure. This reinforces a need to modulate stressors and adequately cope up with stress.

### Self-Medication

In the study of Rahmawati & Bajorek (2017), the various self-medication methods used were examined across the studies the following goals were set for patients with hypertension: (1) to treat their hypertension (e. g. buying captopril without a prescription, eating garlic to lower blood pressure), or (2) control other coexisting health problems (e. g. herbs and painkillers for low back pain,diabetes). Regarding antihypertensive medication self-medication, 11% of people reported buying OTC anti-hypertensive drugs despite being considered a prescription-only medication. A survey of Saudi pharmacists showed that antihypertensive medication (i. e. Unquestioning OTC prescriptions for captopril) were frequently given, despite the fact that there aren’t many articles that have been published that discuss this practice. Findings are crucial because of the potential for improper use of anti-hypertensive medications. The actual percentage of patients buying antihypertensive medication without a prescription may be higher than it is due to the low disclosure of self-medication practices currently reported.

Tan et al. (2017) evaluated hypertensive patients’ perspectives on sensible medication use and hypertension management. In contrast to studies from western and Asian countries, the current findings have successfully explored the various perspectives of appropriate medication use and hypertension management. According to the findings of the current study, key components in the treatment of hypertension with medications include medication adherence and methods to increase medication adherence. Four participants admitted that they occasionally forgot to take their medication. The degree of non-adherence among the participants in the current study could range from skipping one dose of medication per day to skipping several weeks’ worth of medication. This result is consistent with an earlier study on the variation in medication adherence among chronic disease patients in Western nations. The second theme in the current study was self-management of hypertension, which included developing a personal plan and being aware of the symptoms and signs of high blood pressure. In contrast to taking antihypertensive medication, using herbal and traditional therapies was perceived as one of the participants’ personal strategies in controlling blood pressure in the current study and was compared to studies from other countries in Africa, Asia, and Europe.

Furthermore, the study examined the perspectives of hypertensive patients on medication use and emphasized the issue of worries about hypertension management.

Participants’ poor adherence is a result of misinformation regarding the side effects of anti-hypertensive medication. Participants were found to have a lack of understanding of the target blood pressure level, and as a result, there is a low level of blood pressure self-monitoring. It should be promoted at the community level to address the aforementioned gaps through health screening programs and talks given by medical professionals in partnership with NGOs.

According to AlHadlaq et al. (2019), it has been determined that a number of risk factors contribute to hypertension (High Blood Pressure/HBP), a chronic condition that has emerged as a public health issue. HBP is poorly controlled among patients as a result of inadequate care, despite numerous HBP management and behavioral treatment guidelines. The study determined the prevalence of self-management behaviors and investigated the variables influencing self-management behaviors for lowering HBP in hypertensive patients. Gender, age, BMI, the length of HBP, the presence of cardiac disease, and other significant variables have all been linked to behavior, motivation, and self-care confidence. Patients with high blood pressure who visit primary care clinics in the sample population have poor compliance, behavior, motivation, and self-care. Numerous factors were discovered to be connected to bad behavior, lack of luster motivation, and diminished confidence in one’s ability to monitor one’s blood pressure at home, increase exercise, limit salt intake, and value HBP control. It is suggested that health professionals must evaluate patients’ self-care routines and blood pressure control, as well as inform them of the value of HBP monitoring and teach them useful techniques to increase their motivation and confidence and promote better behavior, self-care, and management compliance.

In the study of Lin et al. (2022), 57.8% of Barangay Health Staff (BHS) reported self-medicating, with self-medication of antibiotics accounting for 61.5% of this prevalence. The most frequent causes of self-medication were gastrointestinal symptoms, high blood pressure, a cold, cough, fever, and pains in the head, throat, teeth, muscles, and bones. Painkillers were the most frequently used self-prescribed drug (86.8%), while amoxicillin was the most popular antibiotic (26.5%). Cold and cough remedies, gastrointestinal aids, vitamins and minerals, as well as herbal, anti-hypertensive, contraceptive, hypoglycemic, and vitamin and mineral medications, were additional drug classes used for self-medication. 83.3% of self-medication practices were based on personal experiences, 82.5% on the ease of access to and availability of medications, 41% on academic knowledge of the diseases, 47.4% on treatment options, and 29.5% on the belief that the symptoms were mild. Self-medication was very common among BHS patients. The study demonstrates the need for BHS to provide more in-depth information regarding medication safety, legislation, and appropriate use. The findings of the study advise the Myanmar technical working groups to uphold the national drug policy.

Irwan et al., (2022) revealed that few pertinent studies on patients’ self-care management practices have been conducted in SEA nations where the prevalence of hypertension is rapidly rising. The studies that have been completed show that many hypertensive people have significant behavioral risk factors for heart disease and stroke despite the common medical advice to address these risk factors through self-care management practices. The authors found in their review that a prevalence rate of at least 50% of hypertension people being overweight/obese, having poor physical activity and low-quality dietary intake, and not adhering to medication. Additionally, they discovered that both the facilitators and the inhibitors of putting self-care practices into practice varied, including individual preferences and cultural influences. This information is helpful for nurses because it suggests that when assisting hypertensive patients with managing their self-care and risk reduction practices, nurses working in the community in SEA countries should take cultural considerations into account. Findings show that many of the studies reviewed had high prevalences of obesity, low levels of physical activity, poor dietary intake, and poor adherence to prescribed medications. In some of the reviewed studies, the use of herbal remedies as anti-hypertensive medications is also noted. To better understand individual and contextual influences, more prospective research on the difficulties of self-care management of hypertension is required. Thorough research will be needed to develop tailored solutions to these problems.

### Consultation

The study of Bengtsson et al. (2018) examines the new paradigm of daily self-reporting and self-generation of data by patients in primary care clinical consultation. A mobile phone-based hypertension self-management support system was used with the intention of exploring and describing the structure, topic initiation, and patient contributions in follow-up consultations after eight weeks of self-reporting. By applying BP values to particular circumstances in their own lives, patients’ contributions led to consultations where the patient as a person in context became salient. Additionally, the consultations’ equal participation by patients and medical professionals demonstrated actively involved patients. With a person-centered approach to managing hypertension in primary care, the mobile phone-based self-management support system can be used to support patient involvement in consultations.

Another literature by Drevenhorn, (2018) indicated that blood pressure readings, lifestyle counseling, and translating for doctors are all part of hypertension nursing care. Adapting a new lifestyle for the patient entails taking care of oneself. A middle-range theory of nursing in hypertension care was developed to direct nurses in their practice, in order to improve patient care and design studies for researching nursing in hypertension care, as there is a dearth of research and nursing guidelines. Concepts related to the patient (attitude and beliefs regarding health and illness, autonomy, personality and traits, perceived level of vulnerability, hardiness, sense of coherence, locus of control, self-efficacy, and access to social support and network) and the nurse (applying theories and models for behavioral change in the consultation and using counseling skills, patient advocacy, empowerment, professional knowledge and health education, and support) are presented. Then, Orem’s theory of nursing is incorporated with the concepts related to the consultation (communication, shared decision-making, concordance, coping, adherence, and self-care).

According to Johnson et al. (2021), through a collaborative process known as “shared decision making,” a clinician helps a patient decide what courses of treatment to receive for their health conditions. There are many models of shared decision making, but one recent model that has gained a lot of traction describes a process in which a clinician and a patient communicate about treatment options as well as the patient’s values and preferences before jointly deliberating on a course of action. Patient descriptions and consultation observations of shared decision-making were relatively scant. Frequently, patients had little understanding of their hypertension and were unaware of their treatment options, including the choice to receive none at all. There were few opportunities for patients to learn more about their treatment options and develop a shared understanding with their doctor during consultations. The structured nature of most consultations, which restricted most patients’ contributions to responses to information requests and the distribution of decisions across consultations, limited the opportunities for patients to participate in choice-making. Statin decisions were an exception because many patients understood that they had a choice in this matter. Patients and clinicians agreed that the key decision-makers were the clinicians.

In the study of Świątoniowska-Lonc et al. (2020), patients with hypertension (HT) must constantly check their blood pressure, adhere to therapeutic recommendations, and take care of themselves. A few studies suggest that patient satisfaction with the therapeutic relationship and physician-patient communication may influence the direction and results of the therapeutic process. The relationship between patient satisfaction with physician-patient communication and treatment compliance or self-care in patients with chronic illnesses still needs further study. The purpose of the study was to assess the relationship between self-care and adherence in patients with HT receiving chronic treatment and satisfaction with physician-patient communication. Patients with HT who receive treatment at a specialized clinic express great satisfaction with communication. The physician speaking in an understandable manner and paying attention are the most crucial elements from the patients’ point of view. The most frequent complaints center on the lack of support for question-asking and exclusion from the treatment plan and decision-making. Better self-care and medication adherence in patients with HT are significantly correlated with satisfaction with physician-patient communication. Patients with HT exhibit high levels of self-care competency, particularly in managing one’s own care, and adherence to prescribed medications. Regarding self-maintenance, the lowest self-care quality was discovered.

### Interpersonal Influences Norms

A good health seeking behavior is not easily achieved because it is anchored on a decision-making process that is influenced by individuals or behavior in a household, community norms, and expectations (Haileamlak, 2018). Cislaghi and Heise (2019) stated in their study of using social norms theory for health promotion in low income countries that social norms may hinder or facilitate people’s decision making due to being socially approved by the environment. For women, as housewives, they are under pressure to be healthy so that they will be able to perform their duties. Moreover, it is reported that the constant expectations of the children and the spouse preclude the conduction of health-care seeking behavior. On the other hand, people working outside the home have expressed their experiences of work-related pressures and expectations for maintaining health and treating illness (Morowatisharifabad et al., 2019).

According to Sheleme et al. (2022), contributing factors to hypertensive clients are mainly due to poor medication adherence, lack of information or knowledge about hypertension and frequent changes in regimen are seen with poor blood pressure control. Moreover, a factor which affects hypertensive individuals is a lack of insurance coverage which is considered a critical barrier in getter progressive treatment. Musinguzi et al. (2018) showed how instrumental family members are in the management of hypertension. The results of the study showed that family members did not only offer psychosocial and moral support, but also provided reminders about scheduled appointments, drug refills, and taking medicines. Other sources could also be friends and community members with the same condition, and religious leaders.

### Social Support

Social support or emotional substance can be obtained from interpersonal factors that influence a client’s response to illness, which includes a sense of belongingness or personal involvement in a system or an environment, and social networks. It comes from friends, family members, and even health care providers who provide help when a person encounters problems. To help a client cope with stress and emotion (which are factors that can precipitate high blood pressure), a nurse can assist the client in preparing a list of support people. Moreover, personal health practices like exercise can affect a client’s response to illness. Also, it is an intervention that can help decrease depression and anxiety. When other family members participate in a patient’s exercise, there is an increased social support, improved sense of well-being, and happiness (Videbeck, 2020). On the other hand, Musinguzi et al. (2018) affirmed that patients feel comfortable seeking care from friendly providers.

In terms of social support at a workplace, Yang et al. (2019) stated that to lower presenteeism at work, social support should be improved. Presenteeism refers to loss of productivity at work when employees are not fully functioning due to the presence of illness. Only job stress, which is the most important factor of presenteeism, was looked at as a mediator in the relationship between social support and presenteeism. Also, organizational commitment, which is defined as the degree to which a person identifies with and is involved in a particular organization, plays a substantial mediating role in the relationship between presenteeism and job stress. Social support, a type of intrinsic motivation, boosts organizational commitment and influences the subjective self-worth of strong employees, which reduces presenteeism.

Visvalingam et al. (2019) also argued that maintaining excellent physical and mental health depends heavily on social support, which is the emotional and physical comfort provided to someone in times of need or crisis. Although clever technology devices have contributed to the availability of numerous kinds of social support systems, many people still do not fully understand them. In order to meet the needs of today’s mobile healthcare consumers, several health information systems have been developed, and these support systems are currently available in a variety of formats, including those for tablets and smartphones. It was concluded in the study of Giena et al. (2018) that family and friends should encourage and help the other adults to facilitate changes which will lead to health-promoting behavior.Instances where the family can influence positively is in the preparation of food at home for them.

According to Mo et al. (2022), social support, in its broadest sense, refers to one’s belief that they are supported and that help will be on hand if they need it. The majority of the time, it comes from family, close friends, or the neighborhood. Social network and social capital theories have made an effort to understand the mechanisms underlying how social capital affects health. In particular, social support aids in stress management, offers protective psychological resources (such as self-control, self-efficiency, and mastery), and fosters norms and attitudes that encourage healthy behaviors and results. On the contrary, it is twice as likely to delay receiving health care for people who do not have adequate social support (Reisinger et al., 2018). Most participants in the study of Morowatisharifabad et al. (2019) recognized the family as the most important source of healthcare seeking behavior. Patients who perceived that they had greater social support were more likely to report religious adherence to their antihypertensive medication The study highlighted that the spouse was mainly instrumental, and a woman’s support from her mother or sister was mainly emotional. To improve the quality of care, education among family members about their potential supporting roles is necessary.

One of the autonomous nursing interventions that might help patients and families better manage hypertension is health promotion. Health professionals, particularly nurses, are encouraged to advise senior hypertension patients on health promotion. They must consequently have a thorough awareness of the dietary elements that can influence this illness (Unja et al., 2021). Informational support can be obtained from health care staff, and they have the ability to eliminate ambiguities an individual may have and train self-care towards them. Consequently, other than providing informational and emotional support, they can introduce patients who utilize appropriate therapies as a role model for them (Morowatisharifabad et al., 2019)

### Modeling

Sources of role models may include family role models and peers modeling in taking action for treatment. Participants from the study of Morowatisharifabad et al. (2019) indicated that they sometimes conducted healthcare seeking behavior due to the influence of their friends. A certain participant shared that she has been influenced to protect her health by watching a friend who cares so much about his health and seeks treatment even for the slightest problem in his body. Moreover, relatives who have a history of suffering from a certain similar ailment can serve as another group of role models.

Online platforms can be utilized to manage certain aspects of an individual’s everyday life, including the management of health outside clinical settings. In a study conducted by Bezzubtseva et al. (2022), education through a social media website in hypertension was investigated. Their findings yielded that the most effective method of informing people regarding the primary prevention of hypertension using a social media website is posting a video clip followed by a text post.

The World Health Organization (WHO) recognizes social resources to be a valuable agent for behavior change in the promotion of Health (WHO, 2020). Even with other conditions such as obesity and diabetes mellitus, the use of social media for learning and education has been proven effective (Abedin et al., 2017; Jane et al., 2018). Particularly with the COVID-19 situation, where face-to-face interactions are limited, the use of social media communications and telemedicine has been extensive and useful (Citoni et al., 2022; Kario et al., 2020; Feitosa et al., 2022). Another study conducted by Dhar et al. (2017) has shown that 95% of the participants who joined a Facebook support group for liver transplant patients had a positive impact on their care.

On the other hand, the impact of celebrities on health-related knowledge, attitudes, and behaviors was studied by Hoffman et al. (2017) through a systematic review. It is indicated that celebrities are highly influential people and many of them have used their social status to provide medical advice and/or endorse health products, which is seen today as an increasing trend. Moreover, it is reviewed that celebrities can catalyze herd behavior and provide information that can distinguish endorsed products from others who endorsed it as well. From psychological literature reviews, it is revealed that people are conditioned to react positively to advice given by celebrities and are often pushed subconsciously to follow it. It was suggested that public health authorities can have an opportunity to partner with celebrities, given that people better understand the impact of celebrity involvement.

Overall, the related literature and studies has provided adequate information on the variables that affect the prevalence and management of hypertension among individuals. However, further investigation regarding which health-seeking behavior is highly influenced by interpersonal relationships has yet to be carried out, as well as the factors that can affect health-seeking behavior specific among Filipinos. The Philippines’ health care system is currently developing, and given the restrictions brought about by the COVID-19 pandemic, the investigators would like to explore how hypertensive patients adopted health-seeking behavior to manage their condition at the time.

## Chapter 3 METHODOLOGY

This chapter presents the research method and procedures which will be used in this study. The discussion includes the following sections: (a) research setting, (b) research design, (c) participants and sampling procedure, (d) research instrument, (e) validity and reliability of instruments, (f) scoring procedure, (g) data gathering procedure, and (h) statistical technique. The study will be carried out to determine the health-seeking behaviors and interpersonal influences of the hypertensive patients of an urban barangay in Cagayan de Oro City.

### Research Setting

The study was conducted in a select urban barangay of Cagayan de Oro City, Misamis Oriental, 9000 Philippines. It is situated nearby a creek traversing between two barangays. The investigators were already familiar with the neighborhood since it is where the affiliation for Community Health Nursing was held during the first semester, academic year 2022-2023. The choice of setting was convenient in the data gathering process, for the investigators have already provided nursing care for hypertensive patients there before.

### Research Design

This study has employed the descriptive correlational research design, which utilized quantitative techniques to determine the health-seeking behaviors and interpersonal influences of the hypertensive patients. The design allowed the investigators to obtain information systematically, whether there is a relationship between the independent and dependent variables presented. Using a quantitative approach has enabled the investigators to present data that is objective and can be generalized. According to Bhandari (2022), a correlational research design investigates relationships between variables without the control or manipulation of any of them by the investigators. Furthermore, it reflects the strength and/or direction of the relationship between two or more variables. The direction of the correlation can be described as either positive or negative.

### Participants and Sampling Procedure

The target population of the study were the hypertensive patients residing in an urban barangay in Cagayan de Oro City. The participants were recruited through house-to-house visits by an assigned investigator. Therefore, the data collection was done through the form of home visits. The participants should be clinically diagnosed by a physician with Hypertension, either in a health center or hospital, and/or currently taking anti-hypertensive medications. Also, awareness regarding their own condition was key in selecting the participants. The participants that have been surveyed did not have conditions that resulted from the complications of hypertension, which includes but are not limited to: ischemic stroke, myocardial infarction, and acute kidney failure, for these conditions hinder a patient in adopting health-seeking behaviors, such as exercising. Moreover, the participants chosen were at least 18 years of age, and not suffering from any mental illnesses. The health center of the said barangay was able to provide a census of the known number of patients with hypertension within the site, which is 60. The investigators of the study intended to utilize the universal sampling technique in deciding for the sample size. According to Ramoso (2019), universal sampling technique is where all members of the population will be taken as participants. Therefore, a total of 60 participants will serve as the sample size. According to Adwok (2015), one of the general guidelines for selecting a sample size is to sample the entire population when the number is less than 100. Avron et al. (2019) supported that a universal non-uniform sampling strategy helps achieve optimal complexity for any class of signals.

### Research Instruments

The investigators of this study utilized a modified-survey questionnaire from “The Interpersonal Support Evaluation List (ISEL)” originally developed by Cohen and Hoberman in 1983, which consisted of a list of 40 statements for the general population and 48 for college students regarding the perceived availability of potential social resources (Medical Data Models, 2017). Also, some of the questions were derived from *The Older Adults’ Health Promotion Activity Questionnaire* developed by Chen and Hsieh (2021). It was the key in obtaining significant data, particularly the third part of the questionnaire which included social support, a dimension in Pender’s Health Promotion Model relevant in this study. The modified survey questionnaire was divided into three parts to systematically obtain the variables. The first part of the questionnaire collected the participants’ demographic profiles in terms of their age, sex, civil status, educational level, occupation, monthly income, religion, alcohol use, and tobacco use. The second part determined the participant’s health-seeking behaviors, whether or not they were using alternative medicine, lifestyle modifications, self-medicating, or having consultation with a health care provider. On the contrary, the third and last part of the questionnaire were further divided into three sections; which explored the participant’s three interpersonal influences, which are norms, social support, and modeling, relevant in the management of the patient’s hypertensive condition.

### Validity and Reliability of the Instruments

Validity refers to the accuracy of a method when it measures what it is intended to measure (Middleton, 2019). The research instrument of this study has been validated by three research experts, whose occupations belong to the field of medicine (1) and nursing (2). They provided commentary on the aspects of the instruments that need to be changed and improved.

Moreover, the research instrument was also pilot tested for reliability testing. Reliability refers to how consistently a method measures something (Middleton, 2019). The investigators conducted a pilot testing among 30 hypertensive patients by utilizing google forms, an online tool, where the questions and corresponding scale were encoded. The tool was posted in an online High Blood Pressure Support group in Facebook, a social platform, and was also sent by the investigators to colleagues who have a hypertensive family member. These participants during the pilot testing were not included in the final conduct of the study. This method of pilot testing has been chosen for reasons of safety, convenience, and efficiency.

The survey questionnaire was subjected to a scale reliability test using Cronbach’s Alpha by the university statistician, in order to determine the internal consistency between the variables, signifying whether the instrument was reliable or not. The first domain, which was Norms, obtained a Cronbach’s Alpha of 0.829, which represented the internal consistency as good. Initially, it had 12 items, but 3 items were recommended to be deleted since these variables were unreliable; thus, rendering 9 items as reliable. Next, the second domain was Social Support, which has obtained a Cronbach’s Alpha of 0.783, which also indicated that it was good and acceptable. Initially, it also had 12 items, but 3 items were recommended to be deleted since these variables were unreliable; thus, rendering 9 items as reliable. The last domain was Modeling, in which Cronbach’s Alpha was 0.823, showing that the internal consistency of the variables was also good. It had a total of 12 items, and all of them were interpreted to be reliable. Overall, there were 30 items from the questionnaire that were reliable for data gathering.

### Scoring Procedure

Table 1 shows the scoring guide used in the study. The five-point scale was followed and a range was used to organize responses from the survey questionnaire.

**Table 1.** Scoring Procedure consisting of numerical rating, range, verbal description, and interpretation.

| Numerical Rating | Range | Verbal Description | Interpretation |
| --- | --- | --- | --- |
| 5 | 4.50 – 5.00 | Always | Extremely Influential |
| 4 | 3.50 – 4.49 | Often | Very Influential |
| 3 | 2.50 – 3.49 | Sometimes | Somewhat Influential |
| 2 | 1.50 – 2.49 | Rarely | Slightly Influential |
| 1 | 1.00 – 1.49 | Never | Not Influential at all |

### Data Gathering Procedure

The investigators sought final approval from the school, institutional research ethics board, and the barangay officials. The investigators proceeded in the actual data gathering, which took two days. The investigators chose participants according to the sampling procedure stated in the study. Home visits and interviews were done by the investigators as a means of the recruitment of participants, which lasted for a maximum of 10 to 15 minutes for each visitation. The informed consent form, which served as the cover page of the questionnaire, was read to each participant and obtained their signature prior to administering the research instrument. After the participant has signed the consent form, the investigator proceeded with the actual data gathering while guiding the participants in answering. When the required data has been completely gathered, the investigators organized it in an excel format, which was sent to the university statistician for data analysis and interpretation.

The informed consent is an important component of the study. To thoroughly explain it to the participants, the following components are considered: Providing an overview of the study and its purpose, type of research intervention, participant selection, voluntary participation, procedures, duration of participation, risks, benefits, reimbursements, confidentiality, community considerations and sharing of results, and lastly, the right to refuse or withdraw. It was explained to the participants that the purpose of the study is to determine the health-seeking behaviors resulting from their interpersonal influences to manage their hypertensive condition. Moreover, the research intervention includes requiring the participant to answer a 30-item survey questionnaire from the three main domains: Norms, social support, and modeling. They are to decide a score from 1 (lowest) to 5 (highest) for each item, and that there will be no right or wrong answers. Providing answers is subjective, which means that the participant draws from personal experiences. Also, the demographic profile and health-seeking behaviors were determined. The participants were given two options which they prefer; the investigators can read and explain each item from the questionnaire for them and let them state their answers aloud, or the participants can answer it themselves, with whom enough time will be given.

The investigators also emphasized that the participation is entirely voluntary. If a participant agrees, an informed consent will be provided where the affixation of signature is required. However, the right to withdraw or to refuse at any given time remains valid even after they have signed the informed consent. Withdrawing from the study will not incur any costs. The investigators also have no bearing upon the participant’s decision to withdraw. The data gathered will be returned or disposed accordingly if withdrawal from the study took place before the completion of data collection. The investigators also imposed no pressure of responsibility regarding their choice to participate.

To elaborate the procedure, the survey questionnaires were distributed and collected personally by the investigators, which was done within the participants’ respective homes. In answering the survey, a question may be skipped if a participant wishes to not answer it. The information recorded was kept confidential and contact information such as name, contact number, and address were not recorded in the forms. Only an assigned number by the investigators served as the participant’s identification. Participant involvement lasted only 10 to 15 minutes, or shorter than the indicated time, given that the questions are close-ended.

It was clarified to the participants that there are no anticipated risks related to the conduct of the study. No experiments will be involved or ingestion of substances of any kind. The research study was only limited to interviewing. No data collected will also be shared to other people, as this may violate the participant’s confidentiality. The participants of the study benefited from non-material compensations by the investigators, such as enabling the participants to reflect how good or poor they have been managing their hypertensive condition. It allowed the identification of behaviors that are beneficial or not and let them deal with it accordingly. The investigators also performed a checking of blood pressure using a manual blood pressure apparatus, which has provided the participants their latest blood pressure reading. The benefits from this study are not only limited to the individual, rather it will also contribute at a community level. By participating, the participant contributed to the community by showing whether or not a person’s interpersonal influences has a significant role in promoting health-seeking behaviors, and identify areas of these behaviors that need to be emphasized.

Concerning the reimbursement, the investigators indicated that the participant will not receive any forms of payment that is considered to be more than reimbursements for expenses as a result of being involved in the current study. In maintaining the confidentiality of the gathered data, anonymity was maintained by covering taken pictures of faces during the documentation processes.

### Inclusion and Exclusion Criteria

The inclusion criteria for the selection of participants of the study were that first, the participant should be at least 18 years of age, a Filipino citizen residing at Sitio Calaanan, Barangay Canitoan, being of sound mind, and currently living with significant other/s. The participant must also be aware of their hypertensive condition, as diagnosed either at a Barangay Health Center or hospital, and/or currently taking antihypertensive medication as maintenance that started at any length of time. On the contrary, the exclusion criteria in the selection of participants has identified an individual suffering from the complications of hypertension such as heart attack, stroke, and kidney failure and cognitive impairments or mental disorders such as Alzheimer’s disease, dementia, and schizophrenia. The presence of these conditions had rendered the gathered data unreliable, because it limits the capacity of an individual to practice health-seeking behaviors and provide sound answers. Moreover, those who are living alone were not considered, because the presence of household members was instrumental in promoting health-seeking behaviors, which is also one of the main highlights of the study. The chosen participant did not have a direct relationship with any of the investigators, which has maintained the essence of voluntariness and professionalism. The study involved the elderly; Hence, as a vulnerable group, the investigators evaluated first and asked permission from the significant other present regarding the capability of the elderly to partake in the study.

### Statistical Techniques

In the analysis and interpretation of the data collected, the following statistical measures were employed.

1. To present the demographic profile of the hypertensive patients, the investigators of the study used mean, frequency, and percentage distributions.
2. For problem two, frequency and percentage distribution were utilized in presenting the health-seeking behaviors (alternative management, lifestyle modification, self-medication, and consultation) of the hypertensive patients.
3. For problem three, the frequency, mean, standard deviation, and percentage distribution of the responses were obtained to determine the respective interpersonal influences of the participants.
4. For problem four, Pearson’s Product Moment Correlation Coefficient, also known as Pearson’s Correlation, was used to determine the relationship between the participant’s independent variables (demographic profile and health-seeking behaviors) and dependent variables (interpersonal influences). Particularly, it measures the strength and direction of the association between two variables (Turney, 2022).

## Chapter 4 PRESENTATION, ANALYSIS, AND INTERPRETATION OF DATA

This chapter presents the interpretation and analysis of the data gathered for the study. The presentation follows the logical order of the statement of the problem.

### 1. What is the demographic profile of the hypertensive patients?

Table 2 shows the demographic profile of the participants in terms of age, sex, civil status, religion, and educational level. Among the 60 participants, there were 19 (31.67%) who were aging 65 years old above, 17 (28.33%) aged between 45-54 years old, and 55-64 years old as well, 5 (8.33%) aged from 35-44 years old, and lastly, 1 (1.65%) aged from 18-24 years old and 25-34 years old, which both obtained the lowest response. To highlight the results, most participants aged 65 years old and above, who are individuals diagnosed with hypertension.

**Table 2.** Frequency and percentage distribution of the demographic profile in terms of age, sex, civil status, religion, and educational level. (n=60)

| Variables | Frequency | Percentage |
| --- | --- | --- |
| **Age** |  |  |
| 18-24 years old | 1 | 1.67% |
| 25-34 years old | 1 | 1.67% |
| 35-44 years old | 5 | 8.33% |
| 45-54 years old | 17 | 28.33% |
| 55-64 years old | 17 | 28.33% |
| 65+ years old | 19 | 31.67% |
| **Total** | **60** | **100%** |
| **Sex** |  |  |
| Male | 26 | 43.33% |
| Female | 34 | 56.67% |
| **Total** | **60** | **100%** |
| **Civil Status** |  |  |
| Single | 10 | 16.67% |
| Married | 33 | 55% |
| Separated | 5 | 8.33% |
| Widow/Widower | 12 | 20% |
| **Total** | **60** | **100%** |
| **Religion** |  |  |
| Roman Catholic | 50 | 83.3% |
| Evangelists | 5 | 8.3% |
| Islam | 0 | 0 |
| No Religion | 5 | 8.3% |
| **Total** | **60** | **100%** |
| **Education Level** |  |  |
| Elementary | 11 | 18.3% |
| High School | 27 | 45% |
| Vocational | 1 | 1.7% |
| College | 20 | 33.3% |
| Post-Graduate | 1 | 1.7 |
| **Total** | **60** | **100%** |

Next, among the 60 participants, there were 34 (56.67%) females and 26 (43.33%) males. Most of the participants of this study were female. In terms of civil status, the results showed that among the participants, there were 33 (55%) who are married, 12 (20%) were widow/widower, 10 (16.67%) were single, and lastly 5 (8.33%) are separated. This means that most of the participants are married while least of the participants are separated from their partners.

For religion, the results showed that the highest frequency is 50, which means that 50 of the participants are Roman Catholics, which comprised 83.3%. Two religions have the second highest frequency which is 5, meaning 5 participants were evangelists and another 5 participants have no religion which comprised 8.3% each. The lowest frequency is 0, which means that none of the participants were Islam. To highlight, the results indicated that most of the participants were Roman Catholic.

As for the educational level of the participants, the results showed that the highest frequency is 27, which means that 27 of the participants had finished or reached only high school education, which comprised 45.0%. The second highest frequency is 20, which means 20 participants had a college education level, which comprised 33.3%. The third highest frequency is 11, which means that 11 of the participants had only finished elementary education, which comprised 18.3%. Two educational level categories had the lowest frequency which is 1. This means that 1 participant had a vocational educational level and another 1 participant had a post-graduate educational level, which comprised 1.7% each. Overall, most of the participants had finished high school, while least of the participants had finished vocational and post-graduate educational levels.

Table 3 shows the demographic profile of the participants in terms of occupation, monthly income, alcohol use, and tobacco use. For the occupation, the results showed that the highest frequency is 17, which means that 17 (28.3%) of the participants were unemployed. The second highest frequency is 16, which means 16 (26.7%) participants were self-employed. The third highest frequency is 11, which means that 11 (18.3%) of the participants were homemakers/housewives. The fourth highest frequency is 10, which means that 10 (16.7%) participants were employed. The fifth highest frequency is 5, which means that 5 (8.3%) participants were retired. The second lowest frequency is 1, which means that only 1 participant is a student, which comprised 1.7%. The lowest frequency is 0, which means there are no participants who were unable to work. Overall, the results indicated that most of the participants were unemployed, while none of the participants were unable to work.

**Table 3.** Frequency and percentage distribution of the demographic profile in terms of occupation, monthly income, alcohol use, and tobacco use.

| **Occupation** |  |  |
| --- | --- | --- |
| Variable | Frequency | Percentage |
| Employed | 10 | 16.7% |
| Self-employed | 16 | 26.7% |
| Homemaker/Housewife | 11 | 18.3% |
| Student | 1 | 1.7% |
| Unemployed | 17 | 28.3% |
| Retired | 5 | 8.3% |
| Unable to work | 0 | 0 |
| **Total** | **60** | **100%** |
| **Monthly Income** |  |  |
| Below P10,957 | 46 | 76.67% |
| P10,957 - P21,914 | 9 | 15% |
| P21,914 - P43,828 | 4 | 6.67% |
| P43,828 - P76,669 | 0 | 0 |
| P76,669 - P131,483 | 1 | 1.7% |
| P219,140 and above | 0 | 0 |
| **Total** | **60** | **100%** |
| **Alcohol Use** |  |  |
| Non-alcoholic | 33 | 55% |
| Drinking Alcohol | 27 | 45% |
| **Total** | **60** | **100%** |
| **Tobacco** |  |  |
| Non-smoker | **42** | 70% |
| **Smoker** | **18** | 30% |
| **Total** | **60** | **100%** |

For the monthly income of the participants, the monthly income range that obtained the highest frequency of 46 is below P10,957, which means that there were 46 (76.67%) participants who make this income every month. Next, the monthly income range of P10,957 - P21,914 has obtained the second highest frequency, which is 9. This indicated that 9 (15%) of the participants make this amount every month. Third, only 1 (1.7%) participant makes P76,669 - P131,483 every month. On the other hand, the monthly income ranges of P43,828 - P76,669 & in P219,140 and above obtained 0 responses.

In terms of alcohol use of the participants, among the 60 participants, there were 33 (55%) who were non-alcoholic, while 27 (45%) were alcoholic drinkers.

Consequently, in terms of tobacco use, among the 60 participants, there were 42 (70%) who were non-smokers, while 18 (30%) were tobacco smokers.

### 2. What is the health-seeking behavior among the hypertensive patients?

Table 4 shows the first health-seeking behavior of the participants in the management of hypertension, which is alternative management, such as the use of herbal medicine. The results showed that among the 60 participants, 27 (45%) practiced the use of alternative management. On the other hand, 33 (55%) of the participants did not practice this health-seeking behavior. This outweighed the participants who utilized alternative management.

**Table 4.** Frequency and percentage distribution of the health-seeking behavior, alternative management.

| Variables | Frequency | Percentage |
| --- | --- | --- |
| Practicing | 27 | 45% |
| Not Practicing | 33 | 55% |
| **Total** | **60** | **100%** |

According to Musinguzi et al. (2018), most patients who are seeking complementary medicines used herbs, reporting that they have consulted traditional healers for their hypertensive condition. Patients have considered it as readily accessible and comparably affordable. Thangsuk et al. (2021) also supported that people have the tendency to utilize herbal medication with the belief that it can help make them healthier. However, despite the use of herbal medicine, there is a limited interest in the use of herbal medicine due to the inadequacy of awareness on its impact and usage. Furthermore, reasons for not using herbal medicine included that it is time consuming to prepare; there is insufficient skills among people regarding its preparation; there is lack of easy access to consumption; claims of insufficient efficacy of herbal medicines; and lastly, negative properties of herbal medicine, such that it tastes poorly (Ardakani et al., 2021). Mendoza et al. (2022) stated that most people perceive hypertension as a condition that is treated best by practitioners of biomedical, rather than traditional medicine.

Table 5 shows the second health-seeking behavior of the participants in the management of hypertension, which is lifestyle modification, such as exercising and eating nutritious food. The results showed that among the 60 participants, 39 (65%) practiced lifestyle modification. On the other hand, 21 (35%) of the participants did not practice this health-seeking behavior. The participants who utilized this health-seeking behavior outweighed those who were not.

**Table 5.** Frequency and percentage distribution of the health-seeking behavior, lifestyle modification.

| Variables | Frequency | Percentage |
| --- | --- | --- |
| Practicing | 39 | 65% |
| Not Practicing | 21 | 35% |
| **Total** | **60** | **100%** |

Gabiola et al. (2020) considered lifestyle interventions as first-line therapy for the treatment and prevention of hypertension, and it is gradually appealing in the Philippines, a developing country, where infrastructure, socioeconomic barriers, and resource limitation prohibit access to regular anti-hypertensive medications among patients. An assessment conducted by Bhimla et al. (2017) among Filipinos residing in the Greater Philadelphia Area revealed that only 24.5% of the participants met the aerobic physical activity guidelines, compared to the 49% national average. However, the study of Thomas et al. (2020) among the hypertensive outpatients of Remedios Trinidad Romualdez Hospital concluded that more than 1/3 of the participants followed the diet-related recommendations and 55 out of 80 participants were performing physical exercise, with 24 who were exercising daily for about 16-30 minutes.

Table 6 shows the third health-seeking behavior of the participants in the management of hypertension, which is self-medication, referring to the intake of antihypertensive medication as maintenance or in times of sickness. The results showed that among the 60 participants, 42 (70%) practiced self-medication. On the other hand, 18 (30%) of the participants did not practice this health-seeking behavior. The participants who utilized this health-seeking behavior outweighs those who were not. To support this, the study of Rahmawati and Bajorek (2017) highlighted that there is a high proportion of hypertensive individuals who practice self-medication. Particularly, anti-hypertensive medications are among the 11% of products that are often reported by patients to obtain over-the-counter (OTC) for the purpose of self-medicating. Also, the concurrent use of anti-hypertensive medications together with analgesics and herbal medicine is practiced more often. Moreover, there is an estimated 75% of people with hypertension who reported using anti-hypertensive medication (Samanic et al., 2020). On the contrary, Gutierrez and Sakulbumrungsil (2021) indicated that despite the proven efficacy of anti-hypertensive drugs in the control of blood pressure, there is a low patient adherence, which was reported to be only 20% to 50%.

**Table 6.** Frequency and percentage distribution of the health-seeking behavior, self-medication.

| Variables | Frequency | Percentage |
| --- | --- | --- |
| Practicing | 42 | 70% |
| Not Practicing | 18 | 30% |
| **Total** | **60** | **100%** |

Table 7 shows the fourth health-seeking behavior of the participants in the management of hypertension, which is consultation, referring to having check-ups at a hospital, clinic, or barangay health center. The results showed that among the 60 participants, 49 (81.7%) had their consultation. On the other hand, 11 (18.3%) of the participants did not practice this health-seeking behavior. The participants who utilized this health-seeking behavior outweighed those who were not. Having consultation is the most practiced health-seeking behavior among the participants, followed by self-medication, then lifestyle modification, and alternative management as the least practiced one.

**Table 7.** Frequency and percentage distribution of the health-seeking behavior, consultation.

| Variables | Frequency | Percentage |
| --- | --- | --- |
| Practicing | 49 | 81.7% |
| Not Practicing | 11 | 18.3% |
| **Total** | **60** | **100%** |

Consultation is a significant factor to manage hypertension. According to Johnson et al. (2021), through a collaborative process known as “shared decision making,” a clinician helps a patient decide what courses of treatment to receive for their health conditions. The study of Świątoniowska-Lonc et al. (2020) further supported that patients with hypertension (HT) must constantly check their blood pressure, adhere to therapeutic recommendations, and take care of themselves. However, home-based self-care is the usual first-line strategy of therapy to relieve hypertension-related symptoms, and the decision to finally consult a physician arises when these first-line strategies stop to relieve the symptoms (Mendoza et al., 2022). The current study also revealed that the absence of symptoms and being busy from work served as a barrier in seeking consultation. In the Philippines, most people used local health centers because of the availability of free primary care services and that it is present in every barangay, despite waiting for a long time (Mendoza et al., 2022).

### 3. What are the interpersonal influences among hypertensive patients?

Table 8 shows the first interpersonal influence in the management of hypertension among the participants, which is norms. The variable obtained an overall mean of 3.943 and a standard deviation of 0.814. This indicated that most of the participants often considered the statements stated in the norms, and were interpreted as very influential as they managed their hypertensive condition. To support this, Cislaghi and Heise (2019) stated in their study of using social norms theory for health promotion in low income countries that social norms may hinder or facilitate people’s decision making due to being socially approved by the environment. For women, as housewives, they are under pressure to be healthy so that they will be able to perform their duties. Moreover, it is reported that the constant expectations of the children and the spouse preclude the conduction of health-care seeking behavior.

**Table 8.** Mean and standard deviation distribution of the first interpersonal influence, norms, and the corresponding verbal description and interpretation.

*Norms*
| Indicator | Mean | Standard Deviation | Verbal Description | Interpretation |
| --- | --- | --- | --- | --- |
| 1. I keep myself healthy to be able to take care of my family. | 4.350 | 0.755 | Often | Very Influential |
| 2. I keep myself healthy so that my family will not be worried. | 4.400 | 0.848 | Often | Very Influential |
| 3. I make sure all my family members pay attention to their health. | 4.300 | 0.809 | Often | Very Influential |
| 4. I will not drink alcohol or smoke cigarettes if my family told me so. | 3.583 | 1.418 | Often | Very Influential |
| 5. I find time to exercise. | 3.250 | 1.297 | Sometimes | Somewhat Influential |
| 6. When I feel sick, I voluntarily consult a doctor or take medications. | 3.783 | 1.180 | Often | Very Influential |
| 7. I attend my follow-up check up when advised by my doctor. | 3.950 | 1.126 | Often | Very Influential |
| 8. When prescribed medications, I prefer to seek free medications from the health center. | 3.850 | 1.117 | Often | Very Influential |
| 9. If the health center cannot provide free medications on time, I can ask for help funds from my family or friends to buy my medication. | 4.017 | 1.112 | Often | Very Influential |
| **OVERALL MEAN** | **3.943** | **0.814** | **Often** | **Very Influential** |

To elaborate the results, the second indicator, “I keep myself healthy so that my family will not be worried” has obtained the highest mean, which is 4.40, with a standard deviation of 0.848. This implied that most of the participants often observed this norm, which was considered very influential in the management of their condition. Chacko and Jeemon (2020) claimed that family support in self-care activities is a key factor associated with the control of blood pressure. Joseph et al. (2019) further supported that certain proposed lifestyle changes and self-care strategies are more achievable and sustainable at the same time among individuals with hypertension, if a family-centered approach is employed. Moreover, when patients receive support and encouragement from their community or family, motivations can be greatly improved (Thuy et al., 2021). A participant from the study of Mendoza et al. (2022) also claimed of practicing self-care for the sake of the family, particularly in the consideration of the children’s well-being, in case complications will happen. On the other hand, the fifth indicator, “I find time to exercise” has obtained the lowest mean, which is 3.25, with a standard deviation of 1.297.

This implied that most of the participants only observed this norm sometimes, and was considered somewhat influential in the management of their hypertensive condition. Ghimire et al. (2018) revealed in their study that only one quarter to one third of the Filipino participants had met the recommended exercise guidelines of 150 minutes of moderate physical activity per week. Moreover, Dumlao-Abadilla (2017) wrote that the Philippines, across Asia, is among those with the highest percentage of people who don’t exercise regularly. Most of the reason is due to lack of time, lack of personal motivation, and the distractions that modern life gives. The study of Katanolli et al. (2022) identified two themes which are barriers to physical activity among hypertensive patients; First were structural features such as having crowded sidewalks and lack of green spaces. The second theme was health reasons, such as physical discomfort, stress, and lack of energy. These influence a patient’s decision to manage hypertension.

Table 9 shows the second interpersonal influence in the management of hypertension among the participants, which is social support. The variable obtained an overall mean of 3.674 and a standard deviation of 0.717. This indicated that most of the participants often considered the statements stated in the social support, and interpreted as very influential as they had a social support when it comes to managing hypertension. Mendoza et al. (2022) reviewed that there is a positive association between family social support and hypertension self-care. Also, the study of Giena et al. (2018) supported that family and friends should encourage and help the adults to facilitate changes which will lead to health-promoting behavior. Consequently, it is twice as likely to delay receiving health care for people who do not have adequate social support (Reisinger et al., 2018).

**Table 9.**
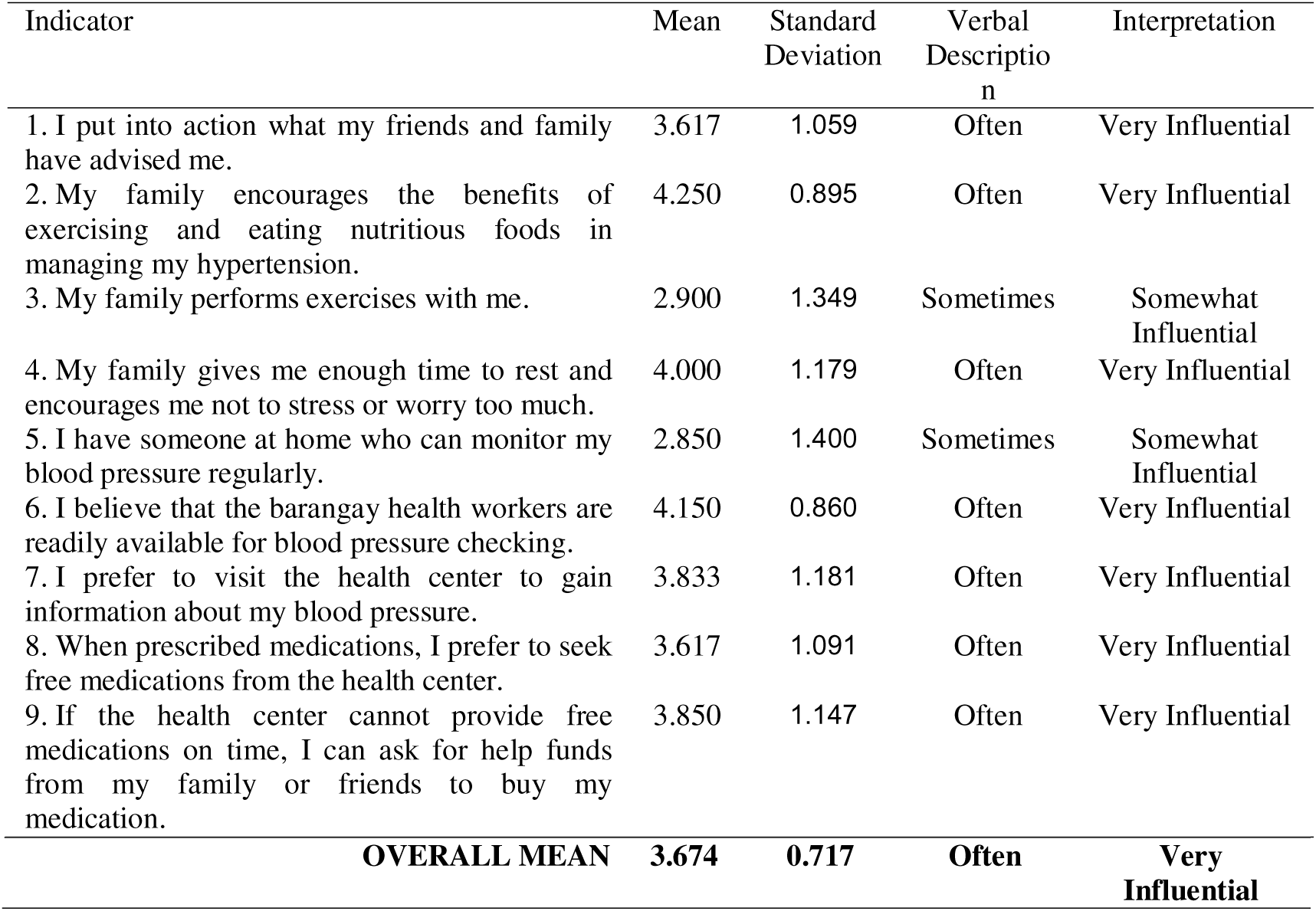
Mean and standard deviation distribution of the second interpersonal influence, social support, and the corresponding verbal description and interpretation.

The second statement, “My family encourages the benefits of exercising and eating nutritious foods in managing my hypertension,” has obtained the highest mean, which is 4.25, with a standard deviation of 0.895 This means that most of the participants were often encouraged by their family to eat healthy food, and to have a healthy lifestyle, which has been very influential in the management of hypertension. Buda et al. (2017) has emphasized that lifestyle modification is the foundation of preventive management among individuals diagnosed with hypertension. In addition, the study of Chacko and Jeemon (2020) found that more than half (53.2%) of men and 62.1% of women from the sample set reported minimal to mild family support regarding self-care activities. On the other hand, moderate to strong family support for self-care activities regarding hypertension management was reported by 47.6% of men and 37.7% of women. It was also discovered that there is an increase in the proportion of individuals with controlled blood pressure when there is an improvement in family support, ranging from minimal (26.2%) to strong support (48.8%) respectively.

On the contrary, the fifth statement, “I have someone at home who can monitor my blood pressure regularly,” has obtained the lowest mean of 2.850, with a standard deviation of 1.400. This showed that most of the participants did not have someone who can monitor their blood pressure and did not also have the blood pressure monitoring equipment. Furthermore, the participants’ blood pressure was only sometimes monitored, and was somewhat influential in the management of hypertension. Sison et al. (2020) highly recognized the importance of home blood pressure monitoring. Furthermore, it was indicated that the reason why hypertension remains a top cause of mortality in the Philippines is due to the lack of awareness, poor compliance, and inadequate blood pressure control. With this, the importance of regular screening in the prevention of hypertension-related complications was stressed by the International Society of Hypertension (ISH). However, barriers to home-based blood pressure monitoring cannot be evaded. Alturi et al. (2023) enumerated several barriers, which included the cost of BP monitors, insufficient patient-provider communication system, and patient health literacy. It was emphasized by doctors that if the cost of BP monitors will be subsidized, most of the other barriers are addressable through patient education and counseling.

Table 10 shows the third interpersonal influence in the management of hypertension among the participants, which is modeling. The variable obtained an overall mean of 3.150 and a standard deviation of 0.770. This indicated that most of the participants sometimes considered the statements stated in the modeling, and interpreted them as somewhat influential as they manage their hypertensive condition.

**Table 10.** Mean and standard deviation distribution of the third interpersonal influence, modeling, and the corresponding verbal description and interpretation.

| *Modeling* |  |  |  |  |
| --- | --- | --- | --- | --- |
| Interpretation | Mean | Standard Deviation | Verbal Description | Interpretation |
| 1. I base my treatment regimen on the experiences of people around me | 3.483 | 1.172 | Sometimes | Somewhat Influential |
| 2. I have a family member or friend with high blood pressure and this encouraged me to practice a healthy lifestyle to manage my condition. | 4.017 | 1.017 | Often | Very Influential |
| 3. I am conscious about my health because I know a friend or relative who died due to a high blood pressure. | 4.167 | 0.977 | Often | Very Influential |
| 4. I seek information about managing high blood pressure from social media, other than the doctor. | 2.950 | 1.254 | Sometimes | Somewhat Influential |
| 5. I join online high blood pressure support groups in social media, such as facebook. | 2.167 | 1.368 | Rarely | Slightly Influential |
| 6. I believe and follow the posts from social media when managing my high blood pressure. | 2.767 | 1.212 | Sometimes | Somewhat Influential |
| 7. I prefer listening to the radio or watching television when obtaining information about managing high blood pressure. | 3.567 | 1.240 | Often | Very Influential |
| 8. I prefer to follow famous people when it comes to eating nutritious foods, exercising, or doing other activities that will help improve my condition. | 2.783 | 1.329 | Sometimes | Somewhat Influential |
| 9. I prefer to take herbal medicine that is advertised by a famous person. | 2.833 | 1.092 | Sometimes | Somewhat Influential |
| 10. I compare myself to others who have high blood pressure, to know if I am managing my condition very well. | 3.100 | 1.189 | Sometimes | Somewhat Influential |
| 11. If the barangay health center will conduct programs and activities for residents with high blood pressure, I will participate. | 3.383 | 1.329 | Sometimes | Somewhat Influential |
| 12. I join groups for yoga, Zumba, or sport sessions to help control my blood pressure. | 2.583 | 1.488 | Sometimes | Somewhat Influential |
| **OVERALL MEAN** | **3.150** | **0.770** | **Sometimes** | **Somewhat Influential** |

The third indicator, “I am conscious about my health because I know a friend or relative who died due to a high blood pressure” has obtained the highest mean, which is 4.16, with a standard deviation of 0.977. This means that most of the participants often observed this modeling, which was very influential in the management of their condition. The statement was supported by Morowatisharifabad et al. (2019) which claims that relatives who have a history of suffering from a certain similar ailment can serve as another group of role models. Li et al. (2021) claims that family history is an important risk factor with significant effect for hypertension. In addition to this, individuals who have family members who suffered from comorbid events such as stroke develop shared chronic management experiences in the family. This resulted in purchasing a blood pressure monitoring device and incorporated self-monitoring in their living, even before being diagnosed with hypertension or having experienced symptoms (Mendoza et al., 2022). Lastly, Anderson et al. (2017) reported that study participants who experienced parental death during childhood had higher systolic BP (SBP) and diastolic BP (DBP) than their unexposed counterparts.

On the contrary, the fifth indicator, “I join online high blood pressure support groups in social media, such as Facebook” has obtained the lowest mean, which is 2.167, with a standard deviation of 1.368. This implied that most of the participants only observed this modeling variable sometimes, and is considered somewhat influential in the management of their hypertensive condition. Fisher et al. (2017) supported that peer support has shown evidence of improving healthy behaviors among patients within under-resourced countries. Moreover, implementing adherence clubs for hypertension control is feasible and can lead to a statistically significant and clinical improvement in self-reported medication adherence which results in BP reduction (Isiguzo et al., 2022). However, there is a barrier in access to technology, considering that most of the participants of this study came from low-income households, and that most of the participants are elderly. A survey conducted by the Pew Research Center has revealed a considerable gap in the internet usage among older and younger Filipinos, and particularly those with low and high educational attainment. The use of social media is more prevalent among younger people than older people, and 61% of low-income individuals use the internet, compared to the ones coming from high-income brackets, which is 80%. In contrast, Mancheno et al. (2021) found that encouraging patients who have poorly controlled hypertension to tweet or retweet health content on twitter did not show significance in improving systolic blood pressure and patient activation measure (PAM) within 6 months.

### 4. Is there a significant relationship between the interpersonal influences, health-seeking behavior, and demographic profile among hypertensive patients?

Table 11 shows the significance of the demographic profile of the participants, an independent variable, towards the participants’ interpersonal influence, a dependent variable, using Pearson’s chi-square tests. It showed that the variables-sex, civil status, religion, education level, occupation, and tobacco use had a p-value of greater than the 0.05 level of significance. It implied that these variables did not have a significant relationship with the interpersonal influences of the participants. According to LaMorte (2019), a p-value greater than 0.05 is interpreted as not statistically significant, which means that there is insufficient strong evidence to reject the null hypothesis. However, this does not indicate that the aforementioned variables are not important at all; rather, there is only no association or good evidence that these variables are factors that can affect how the participants respond to the influences of their interpersonal relationships when it comes to adopting health-seeking behaviors.

**Table 11.** Pearson correlation distribution between the demographic profile and interpersonal influences of the hypertensive patients. (n=58)

| Variable | Pearson Correlation | P-value* | Interpretation |
| --- | --- | --- | --- |
| Age | 43.115 | 0.000 | Significant |
| Sex | 7.487 | 0.058 | Not Significant |
| Civil Status | 10.076 | 0.344 | Not Significant |
| Religion | 11.471 | 0.075 | Not Significant |
| Education Level | 7.219 | 0.843 | Not Significant |
| Occupation | 18.394 | 0.243 | Not Significant |
| Monthly Income | 17.550 | 0.041 | Significant |
| Alcohol Use | 8.765 | 0.033 | Significant |
| Tobacco Use | 5.455 | 0.141 | Not Significant |

*Legend:* \*- Correlation is significant at the 0.05 level

On the other hand, there were three variables from the demographic profile that had a p-value below the 0.05 level of significance. These are age, which has a p-value of 0.000; monthly income, with a p-value of 0.041; and lastly, alcohol use with a p-value of 0.033, respectively. This implied that these variables had a significant relationship with the interpersonal influences of the participants, therefore rejecting the null hypothesis. These variables affected how hypertensive patients responded to their interpersonal influences in adopting health-seeking behavior to manage hypertension. According to Chen and Hsieh (2021), age and self-efficacy had relatively higher contributions toward participation in health promotion activities. Bernard and Tolentino (2019) claimed that the top health-seeking behavior among the elderly is self-care, which includes eating well, exercising, and having fun, but it should be mentioned that elderly people prefer to practice better self-care if they have strong relationships and participate in social activities. Despite the fact that elderly people rarely seek medical attention, the findings suggest that as social support grows, so does the frequency of the aforementioned health-seeking behavior. There is a difference in the prevalence of treatment-seeking behavior for hypertension among younger and older individuals. The level of awareness was better among the participants from the age group of 20-29 (57%), compared to those belonging to 30-39 years old which comprised only 47.5%. Moreover, only 19.5% among these self-reported participants have sought treatment and are currently on medications (Krishnamoorthy et al., 2022). Younger participants from the study of Mendoza et al. (2022) stated that regular medication may not be imperative, reasoning that they are capable of withstanding the symptoms of hypertension.

One’s financial resources is a significant determinant of adopting health-seeking behavior, when advised by families, friends, or health care workers. With this, the significance of monthly income was supported by Musinguzi et al. (2018) who stated that patients with a greater economic resources are likely deemed to seek treatment from doctors in private facilities and spend a more considerate amount, compared to those belonging to a low socioeconomic index. Hogg (2022) added that impoverished populations might face more perceived barriers than general populations. Someone facing impoverishment is likely to use their financial resources to meet basic needs such as food, water, and clothing before purchasing medication, particularly when they have little understanding of the importance of the medication and complications of hypertension. Upon the diagnosis of hypertension, those who belong in low socioeconomic status are less likely to afford out-of-pocket expenses to purchase antihypertensive medication, which can eventually lead to uncontrolled hypertension (Schutte et al., 2021). Alcohol use, consequently, has also shown significance on the adoption of health-seeking behavior based on a patient’s interpersonal influences. Li and Sun (2022) found that adults who regularly drank alcohol, compared to nondrinkers, had a decreased chance of seeking health care. Furthermore, if there is a higher willingness to bear risk will decrease the probability of seeking health care (Lutter et al., 2019). Kim et al. (2018), lifestyle behaviors like smoking, drinking, and physical inactivity are associated with poor health conditions. This in turn is more likely to cause negative experiences with seeking health care and become less satisfied with it.

Table 12 shows the significance of the health-seeking behavior towards the participants’ interpersonal influences, also using Pearson’s chi-square tests. Among the four, two variables had p-values of greater than the 0.05 level of significance. These variables were alternative management, with a p-value of 0.100, and consultation with a p-value of 0.092. It implied that these variables did not have a significant relationship with the interpersonal influences of the participants. Consequently, the null hypothesis cannot be rejected. Alternative management and consultation were the health-seeking behavior practices that were insignificantly influenced by a patient’s interpersonal relationships in managing hypertension. It did not indicate that these variables are unimportant; rather, it functions independently from a patient’s interpersonal influences.

**Table 12.** Pearson correlation between the health-seeking behavior and interpersonal influences among hypertensive patients. (n = 58)

| Variable | Pearson Correlation | P-value* | Interpretation |
| --- | --- | --- | --- |
| Alternative Management | 6.248 | 0.100 | Not Significant |
| Lifestyle Modification | 13.454 | 0.004 | Significant |
| Self-Medication | 13.647 | 0.003 | Significant |
| Consultation | 6.435 | 0.092 | Not Significant |

*Legend:* \*- Correlation is significant at the 0.05 level

On the other hand, two variables had p-values below 0.05; these were lifestyle modification, which had a p-value of 0.004, and self-medication, with a p-value of 0.003, respectively. This means that these variables had a significant relationship with the interpersonal influences of the participants, thereby rejecting the null hypothesis. Both lifestyle modification and self-medication were the health-seeking behavior practices that were significantly influenced by the participants’ interpersonal relationships.

Hypertension necessitates patients and their respective families to participate actively in their self-care, which involves activities like taking medication, becoming aware of complications of the disease and danger signs, having follow-up regularly, and most importantly, making lifestyle changes and managing changes in emotion (Gupta et al., 2020). Irwan et al. (2022) indicated that lifestyle modifications in diet, exercise, and stress management are effective to reduce the incidence of and improve management of hypertension. When combined with improved adherence to medications, these behaviors represent a cornerstone of recommended care for hypertension care (Carey et al., 2019). However, Giena et al. (2018) explained that extensive efforts are needed as influenced by one’s social context. Mendoza et al. (2022) indicated that social circles are primary sources of information for various treatments for hypertension, such as the types of food, herbal supplements, and medications that contribute good outcomes for hypertension. Furthermore, family members and social networks had a significant role in the therapeutic itinerary for hypertension. A good instance would include reminding participants about the intake of medication, collecting free medicine from health centers on behalf of them, and giving encouragement to seek health care or pursue self-care. Familial affection drives a person’s motivation to start adhering to prescribed treatment regimen. This puts emphasis that the care process is not produced by individuals merely for their own well-being but rather how it is relational in nature and created with and against significant others. In contrast, the same study highlighted that medicine stock-outs in health centers posed financial burdens among patients, because free medications that are supposed to be collected monthly were not always available. This in turn could result in intermittent adherence.

## Chapter 5 SUMMARY, CONCLUSIONS, AND RECOMMENDATIONS

This chapter presents the summary of the results, conclusions, and the investigators’ recommendations based on the results of the study.

### Summary

The study was implemented to determine the health-seeking behavior and interpersonal influences among the hypertensive patients of Barangay Canitoan, Cagayan de Oro City. The study was anchored on the Health Promotion Model (HPM) of Nola J. Pender (1982).

Two factors fall under the independent variable; first was the demographic profile of the participants which included: age, sex, civil status, education level, occupation, monthly income, religion, alcohol use, and tobacco use. Second was the health-seeking behavior among the participants, which were alternative management (use of traditional medicine), lifestyle modification, self-medication, and consultation. The dependent variables were the interpersonal influences of the participants in terms of norms, social support, and modeling. Then, the investigators sought to identify if there is a significant relationship between the interpersonal influences, versus the health-seeking behavior and demographic profile of the participants.

A descriptive correlational research design was utilized while employing a quantitative approach to present an objective and generalized data. The study consisted of 60 hypertensive patients residing at Barangay Canitoan, Cagayan de Oro City. The sample size was based on the census provided by the health center of Sitio Calaanan; thus, universal sampling was employed. The research instrument has been tested for reliability among 30 hypertensive patients. Consequently, data collection was done through the administration of a modified survey questionnaire. Data was analyzed using descriptive statistics, and Pearson’s correlation for determining the significance of the independent variables towards the dependent variable.

### Conclusion

1. Based on the demographic profile, the study yielded that 31.67% of the participants belong to the age group of 65 and above. The participants were 56.67% female and 43.33% male. Moreover, 55% were married, and 83.3% identified as Roman Catholics. In terms of education level, 45% of the participants finished only high school education. The occupation status showed that 43.33% are currently working, 46.67% were unemployed, 8.3% were retired, and there is only one student which accounted for 1.7%. Moreover, based on socioeconomic status, 76.67% reported a monthly income bracket of below P10,957, which is interpreted as poor according to the Philippine Institute for Development Studies (2018). The results also showed that 27% were alcoholic while 33% were not. Consequently, 18% were smoking tobacco while 42% were non-smoker.
2. As for the health-seeking behavior, most of the participants sought healthcare provider consultations (81.7%), followed by self-medication (70%), lifestyle modification (65%), and the least was alternative management (45%).
3. The participants consider the interpersonal influence, norms, as very influential in managing their condition, for the indicators under the said variable are often practiced. The indicator often observed and considered very influential by the participants was “I keep myself healthy so that my family will not be worried.” This implies that they practiced self-care to not have their families worried over them. On the contrary, the indicator, “I find time to exercise” was only practiced by the participants sometimes and was considered somewhat influential. This means that the participants only exercise sometimes.
4. The participants considered social support as very influential to their hypertension management. Most of them indicated that, “My family encourages the benefits of exercising and eating nutritious foods in managing my hypertension.” This means that most of the participants are being encouraged by their family members to eat healthy foods, and to have a healthy lifestyle, which has been very influential in hypertension management. On the other hand, minority of the participants indicated that, “I have someone at home who can monitor my blood pressure regularly.” This shows that the participants do not have someone who can monitor their blood pressure and does not also have the blood pressure monitoring equipment.
5. For modeling, the participants only consider it as somewhat influential in managing their condition, and that the indicators are observed or practiced only sometimes. Most of them indicated that, “I am conscious about my health because I know a friend or relative who died due to a high blood pressure.” This means that most of the participants keep themselves healthy more often because of a known history of mortality within the social circle due to hypertension. On the other hand, the participants only sometimes observed the indicator, “I join online high blood pressure support groups in social media, such as Facebook.” This implies that most of the participants do not join online support groups for hypertension, and was only considered somewhat influential in hypertension management.
6. Lastly, there were three (3) variables among the nine identified variables under the demographic profile that showed significant relationships between the interpersonal influences, namely: age, monthly income, and alcohol use. Meanwhile, there were two (2) out of four variables categorized under health seeking behaviors that are significantly associated with interpersonal influences, namely: lifestyle modification and self-medication.

The findings of this study provide important implications in the adoption of health-seeking behavior among hypertensive patients as influenced inter-personally by their families, friends, peers, and even health care workers. It was found that lifestyle modification and self-medication as preventive and therapeutic measures for hypertension was significant and very influential among the participants. In addition, it has a significant relationship with the interpersonal influences; which includes norms, social support, and modeling. This implies that when these practices are reinforced by an individual’s significant other, the adoption of health-seeking behavior is likely to succeed. To highlight, enforcing the health-seeking behavior through setting expectations (norms) and providing emotional and instrumental support (social support) can be very influential in hypertension management.

The direct involvement of significant others can efficiently produce positive outcomes. Additionally, there was low prevalence of the use of alternative medicine among the participants, thereby resulting in an insignificant association with one’s interpersonal influences. Consultation, on the contrary, despite showing the highest prevalence of being practiced, appears independent from interpersonal influences. This indicates that the choice to consult with a professional for the diagnosis of hypertension or follow-up check-up is an intrapersonal perspective. Moreover, an individual’s demographic profile plays a key role in adopting health-seeking behavior, which includes age, monthly income, and behavioral practices like alcohol use. These factors have an important bearing on whether an individual will put into action the healthy recommendations given by significant others to manage hypertension or not.

### Recommendations

1. The efficacy of reinforcing health-seeking behavior for hypertension among patients patients can be improved by health workers through the provision of health teachings that not only pertains to hypertension itself, but also emphasizes the active role of an individual’s interpersonal influences such as family members, friends, and even peers in managing the disease. The direct involvement of significant others can produce positive behavioral outcomes, and the findings of this study necessitates their inclusion.
2. Significant others can help enforce healthy practices among hypertensive members of the family, particularly when health expectations (norms) are set, and emotional and instrumental support (social support) are adequately provided. Based on the findings, observing lifestyle modifications and medicating with anti-hypertensive drugs are areas considered very influential, and family members should give recognition towards these.
3. The investigators recommend other studies to look into the varying hypertension status of patients and its relationship with adopting health-seeking behaviors, which this study has not further looked into. Moreover, future investigators many conduct further studies on the components of Pender’s Health Promotion Model, such as exploring the association between one’s interpersonal influences and situational influences.

## Data Availability

All data produced in the present study are available upon reasonable request to the authors.

